# Predicting Type 2 Diabetes Metabolic Phenotypes Using Continuous Glucose Monitoring and a Machine Learning Framework

**DOI:** 10.1101/2024.07.20.24310737

**Authors:** Ahmed A. Metwally, Dalia Perelman, Heyjun Park, Yue Wu, Alokkumar Jha, Seth Sharp, Alessandra Celli, Ekrem Ayhan, Fahim Abbasi, Anna L Gloyn, Tracey McLaughlin, Michael Snyder

**Affiliations:** Department of Genetics, Stanford University, Stanford, CA 94305, USA; Department of Medicine, Stanford University, Stanford, CA 94305, USA; Department of Pediatrics, Stanford University, Stanford, CA 94305, USA; Stanford Diabetes Research Centre, Stanford University, Stanford, CA 94305, USA

**Author notes:** These authors contributed equally (Tracey McLaughlin, Michael Snyder).

**Keywords:** T2D, prediabetes, CGM, machine learning, time-series, phenotype, insulin resistance, heterogeneity, precision medicine, incretin effect, β-cell function, metabolism, OGTT

## Abstract

Type 2 diabetes (T2D) and prediabetes are classically defined by the level of fasting glucose or surrogates such as hemoglobin HbA1c. This classification does not take into account the heterogeneity in the pathophysiology of glucose dysregulation, the identification of which could inform targeted approaches to diabetes treatment and prevention and/or predict clinical outcomes. We performed gold-standard metabolic tests in a cohort of individuals with early glucose dysregulation and quantified four distinct metabolic subphenotypes known to contribute to glucose dysregulation and T2D: muscle insulin resistance, β-cell dysfunction, impaired incretin action, and hepatic insulin resistance. We revealed substantial inter-individual heterogeneity, with 34% of individuals exhibiting dominance or co-dominance in muscle and/or liver IR, and 40% exhibiting dominance or co-dominance in β-cell and/or incretin deficiency. Further, with a frequently-sampled oral glucose tolerance test (OGTT), we developed a novel machine learning framework to predict metabolic subphenotypes using features from the dynamic patterns of the glucose time-series (“shape of the glucose curve”). The glucose time-series features identified insulin resistance, β-cell deficiency, and incretin defect with auROCs of 95%, 89%, and 88%, respectively. These figures are superior to currently-used estimates. The prediction of muscle insulin resistance and β-cell deficiency were validated using an independent cohort. We then tested the ability of glucose curves generated by a continuous glucose monitor (CGM) worn during at-home OGTTs to predict insulin resistance and β-cell deficiency, yielding auROC of 88% and 84%, respectively. We thus demonstrate that the prediabetic state is characterized by metabolic heterogeneity, which can be defined by the shape of the glucose curve during standardized OGTT, performed in a clinical research unit or at-home setting using CGM. The use of at-home CGM to identify muscle insulin resistance and β-cell deficiency constitutes a practical and scalable method by which to risk stratify individuals with early glucose dysregulation and inform targeted treatment to prevent T2D.

**Article Highlights:**

1. The study challenges the conventional classification of type 2 diabetes (T2D) and prediabetes based solely on glycemic levels. Instead, the results highlight the heterogeneity of underlying physiological processes that represent separate pathways to hyperglycemia. Individuals with normoglycemia and prediabetes can be classified according to the relative contribution of four distinct metabolic subphenotypes: insulin resistance, muscle and hepatic, β-cell dysfunction, and incretin defect, which comprise a single dominant or codominant physiologic process in all but 9% of individuals.
2. Use of multiple time points during OGTT generates time-series data to better define the shape of the glucose curve: the application of a novel machine learning framework utilizing features derived from dynamic patterns in glucose time-series data demonstrates high predictive accuracy for identifying metabolic subphenotypes as measured by gold-standard tests in the clinical research unit. This method predicts insulin resistance, β-cell deficiency, and incretin defect better than currently-used estimates, with auROCs of 95%, 89%, and 88%, respectively.
3. The muscle insulin resistance and β-cell deficiency prediction models above were validated with an independent cohort and then tested using glucose data series derived from OGTT performed at home with a continuous glucose monitor (auROC of at-home prediction of insulin resistance and β-cell deficiency is 88% and 84%, respectively). This approach offers a practical and scalable method for metabolic subphenotyping and risk stratification in individuals with normoglycemia or prediabetes, with potential to inform targeted treatments to prevent progression to T2D.

## Introduction

Type 2 diabetes (T2D) affects over 537 M adults globally^1^. Diabetes and prediabetes are defined by measures of glucose elevation, but the underlying physiology is complex and differs between individuals. It is traditionally suggested that individuals who develop T2D have both insulin resistance (IR) and β-cell dysfunction. However, we and others have shown that individuals with prediabetes and early T2D exhibit varying degrees of insulin resistance and β-cell dysfunction^2,3^, as well as defects in incretin action^4^ and hepatic glucose regulation^5^. These physiologic underpinnings of T2D are present before the onset of overt hyperglycemia^6^, but they are not easy to identify outside of specialized metabolic research units. In particular, insulin resistance (IR) would be useful to detect in stages prior to overt hyperglycemia because it is highly prevalent^7^ and increases risk not only for T2D, but also for cardiovascular disease (CVD), stroke, hypertension, non-alcoholic liver disease, and cancer^7–11^, and is largely reversible with lifestyle interventions such as weight loss and physical activity^12,13^.

We propose that classifying individuals with prediabetes, diabetes risk factors, and newly diagnosed T2D according to their underlying metabolic physiology rather than level of glycemia is possible and could enable a precision medicine approach to diabetes prevention and treatment. Not only might metabolic subphenotypes pose differential risk for conversion to T2D, diabetes-related complications, and/or CVD, they might determine relative efficacy of pharmacologic and lifestyle therapies. Multiple approved T2D treatments targeting specific physiologic components of glucose regulation now exist, virtually all of which have also been proven to prevent progression to T2D in high-risk individuals^14–17^. Efficacy in diabetes prevention varies between individuals however, with more obese and younger individuals responding better to metformin over lifestyle, and older less obese individuals responding better to lifestyle^14^. Similarly, among non-insulin treated diabetics starting monotherapy for T2D, females and patients with higher BMI responded better to thiazolidenediones, whereas males and patients with lower BMI responded better to sulfonylureas^18^. Heterogeneity in responses may reflect the heterogeneity in underlying physiology, since both pharmacologic and lifestyle interventions target different physiologic pathways to hyperglycemia. Lifestyle interventions such as weight loss and exercise reduce insulin resistance^18,19^, whereas reducing dietary sugar and glycemic load might benefit those with β-cell deficiency and/or incretin deficits; metformin reduces hepatic glucose production, acarbose mimics prevents absorption of dietary carbohydrates in the small intestine, thiazolidenediones are powerful insulin sensitizers, and GLP-1 agonists augment β-cell insulin secretion and reverse insulin resistance via weight loss. Thus, phenotyping individuals according to underlying metabolic physiology might allow for targeted therapy at an early stage to prevent and/or treat T2D.

There is growing interest in identifying scalable cost-effective tests which capture the complexity of T2D phenotypes in order to inform precision medicine approaches. These include partitioned genetic risk scores^20^, clinical and demographic features^21^, glucose metabolism biomarkers, clinical outcomes^22–27^, and even wearable devices^2,28^. The most common test for glycemic disorders is the oral glucose tolerance test (OGTT), which has been standardized and used globally for over 100 years. Limitations of fasting plasma glucose in identifying early dysglycemic states led to widespread use of the OGTT for epidemiological studies as well as clinical practice. Metrics such as the 2 hour glucose, 1 hour glucose, area under the curve, and shape of the curve during a 3 or 5 timepoint OGTT have shown utility in predicting progression to T2D, development of microvascular disease, cardiovascular events, and total mortality^29,30^. Furthermore, differences in the relationship between fasting and 2 hour glucose elevations with hepatic IR, muscle IR, and β-cell function highlighted the potential for OGTT to define underlying pathophysiology^29,31–36^. The burden of performing this test has limited its use clinically, particularly with regards to multi-time point analyses, which in 1980 were simplified to the fasting and 2 hour time points alone. With the advent of CGM, it is possible to perform facile time-series measurements of glucose levels and OGTTs. We hypothesized that features of a 16-point glucose curve generated in response to standardized administration of an oral glucose load could identify the specific metabolic abnormalities underlying glucose dysregulation, which, if replicated by continuous glucose monitor (CGM) derived curves, would represent a scalable, practical, and relatively cost-effective method that could facilitate individualized prevention and treatment of T2D, thereby enhancing precision approaches to diabetes.

In this work, we quantify the degree to which the physiologic basis for glucose dysregulation differs between individuals, who can be classified according to their metabolic subphenotypes. We further present a novel and comprehensive framework using machine learning by which metabolic subphenotypes can be identified using features of the glucose time-series extracted during a 16-point glucose tolerance test. We then demonstrate that this can be performed at-home using a CGM. The accuracy of this method, both in plasma and CGM, to identify underlying metabolic dysfunction is higher than standard measures of hyperglycemia (e.g., fasting glucose or HbA1c), existing biomarkers of metabolic disease (e.g., HOMA), and genetic risk score. Together these results demonstrate the full potential of glucose measurements by CGM to define the pathophysiology underlying early dysglycemic states.

## Results

### Overview of the study and cohorts studied

We enrolled 56 individuals without history of diabetes and fasting plasma glucose <126 mg/dL, who were classified as having normoglycemia (n = 33) or prediabetes (n = 21), as well as two with T2D\ according to American Diabetes Association HbA1c criteria (<5.7%, 5.7-6.4%, ≥ 6.5%). We then used a machine learning approach to determine if we could distinguish metabolic subphenotypes based on the shape of glucose curves from a 16-point OGTT performed in the Stanford University School of Medicine’s clinical translational research unit (CTRU) (**Figure 1A**) as well as the average of two OGTTs performed at home using a CGM (**Figure 5A**). Metabolic physiology was characterized using gold-standard tests as described below.

**Figure 1:**
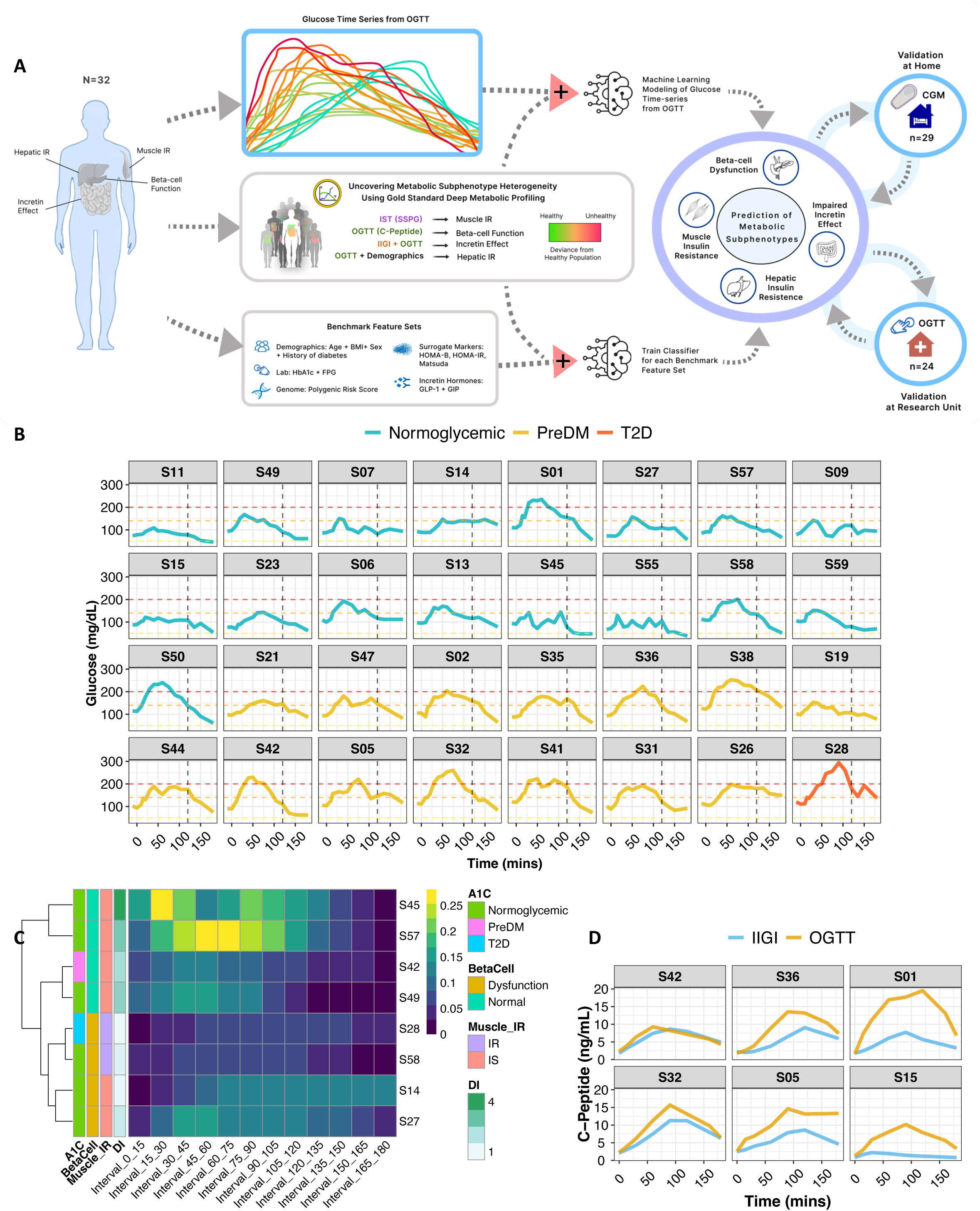
Metabolic subphenotyping study overview. **(A)** Study design. **(B)** Heterogeneity in glucose time-series following a frequently sampled oral glucose tolerance test (OGTT). Time=0 on the x-axis represents the time when participants start drinking the glucose drink. The vertical black dashed line represents the time at 120 minutes where the glucose level is measured for clinical diagnosis of diabetes. Glucose value above 200 mg/dL (red dashed line) represents a diabetes status, glucose value below 140 mg/dL (orange dashed line) represents a normoglycemic status, glucose value between 140-200 mg/dL represents a prediabetes status, and glucose value below 70 mg/dL (yellow dashed line) represents a hypoglycemia status. **(C)** Eight examples of insulin secretion rate (C-peptide deconvolution) normalized by insulin resistance (SSPG) used to infer β-cell function. **(D)** Illustration of incretin effect variability. Six examples of C-peptide concentration during OGTT and IIGI. The incretin effect is calculated as the area between the C-peptide OGTT and IIGI divided by the area under C-peptide at OGTT. Left panel indicates poor incretin response (S42 and S32), middle panel indicates moderate incretin response (S38 and S05), and the right panel indicates robust incretin response (S01 and S15).

Three cohorts were included in this study: (1) an initial cohort for training (n=32) and testing the model, (2) a validation cohort (n=24), (3) an at-home CGM cohort (n=29). For the main training cohort, thirty-six participants were enrolled, and thirty-two completed all metabolic tests in the CTRU. These 32 participants were included in the final analyses as the initial training study cohort. An independent validation cohort of 24 participants was recruited separately and analyzed. Finally, to test the feasibility of home CGM for metabolic subphenotyping, 29 participants (5 of the initial cohort and 24 of the validation cohort) participated in the at-home OGTT/CGM study, and completed a minimum of two metabolic tests in the CTRU. This cohort also enabled us to compare concordance of two home CGMs as well as home CGM versus CTRU CGM and CTRU plasma values during OGTT. The protocol was approved by the Stanford Internal Review Board and conducted according to the principles of the Declaration of Helsinki and Good Clinical Practice (**Methods**). The cohorts’ characteristics are summarized in **Table 1** and **Supplementary Table S1**, and were well matched with an average age of 55 yrs, BMI 26 kg/m2, relatively equal ratio of male/female sex, 74% Caucasian and 27% Asian ethnicity, and HbA1c of 5.6%.

### Unique glycemic responses to oral glucose load

To characterize the dynamic pattern of the glucose time-series during the oral glucose tolerance test (OGTT), we measured plasma glucose concentrations at 5-15 min intervals (16 time points) for 180 min following administration of a 75g oral glucose load under highly standardized conditions (see Methods) in the Stanford CTRU. We then performed deep metabolic profiling using gold-standard quantitative tests with the goal of assessing four distinct physiologic phenotypes known to contribute to glucose dysregulation and T2D: muscle insulin resistance (IR), β-cell dysfunction, impaired incretin action, and hepatic IR (**Figure 1A, Methods**). We developed a machine learning algorithm that used the glucose time-series to predict metabolic subphenotypes (**Figure 1A**).

As shown in **Figure 1B**, OGTT glucose time-series profiles were very heterogeneous across individuals, despite similar classification based on OGTT 2 hour plasma glucose levels (normoglycemic: glucose<140 mg/dL, prediabetes: 140≤ glucose <200 mg/dL, diabetes: glucose ≥200 mg/dL). The shape of the curve was remarkably different with regard to ascending and descending slopes, peak height, and number of peaks, among other characteristics. As exemplified by participants S28, S42, and S01, some participants exhibited large rapid glucose spikes, whereas others (S21, S23) demonstrated little excursion at all. Some exhibited multiple glucose spikes (S42, S47, S55) whereas others had only a single spike (S32). Additional differences are apparent in **Figure 1B**.

### Heterogeneous responses to metabolic testing

Muscle insulin resistance was measured by the modified insulin-suppression test (IST) and expressed as steady-state plasma glucose (SSPG)^37–39^. Participants were categorized as insulin sensitive (IS) if SSPG was <120 mg/dL and insulin resistant (IR) if their SSPG was ≥120 mg/dL. Our designation of IR encompasses individuals above the 50% of the distribution of SSPG among 490 healthy volunteers as previously described^40^. It includes those with “moderate” elevations in SSPG who are more accurately defined as “non-IS”. This value also represented a natural cut-point in the distribution pattern of our study cohort (**Supplementary Figure S1A**), and is clinically attractive as individuals with SSPG > 120 mg/dL, even if moderately IR, respond to both lifestyle and pharmacologic treatments that target IR. As previously observed^40^, SSPG values spanned a wide range from 40 (subject S19) to 278 mg/dL (subject S38) (**Figure 2A, Supplementary Table S2**). Importantly, in the current study, SSPG values indicating IR (>120 mg/dL) or IS (<120 mg/dL) varied despite glycemic status. For example, some participants with elevated (prediabetes range) HbA1c, had low SSPG values indicating insulin sensitivity, whereas others with normoglycemia were quite insulin resistant (examples in **Figure 2A**). These data emphasize that insulin resistance is not synonymous with glycemic elevations.

**Figure 2:**
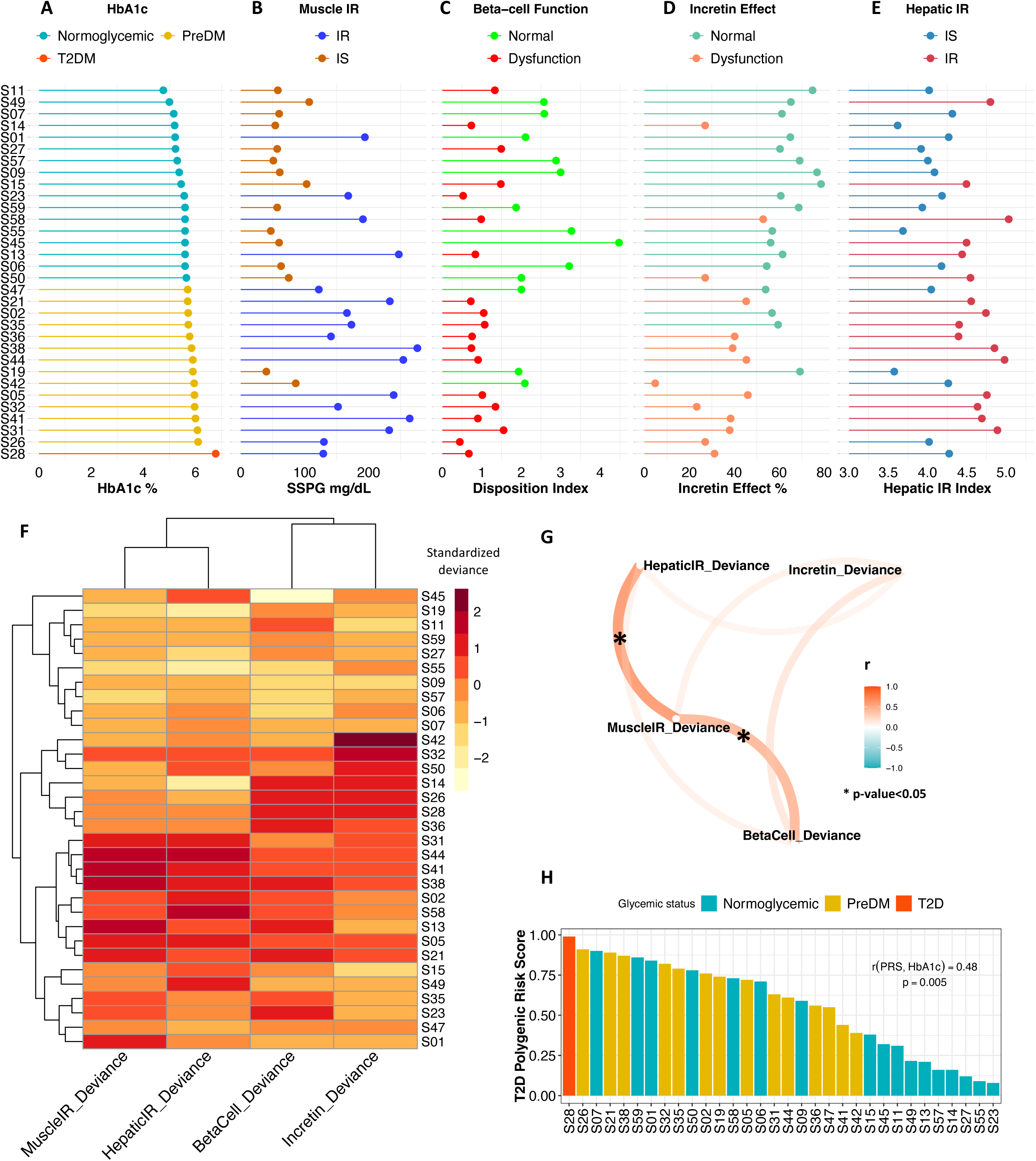
Heterogeneity in metabolism and determination of the dominant metabolic subphenotype. Detailed illustration of the four metabolic measures for our cohort sorted by the HbA1c **(A)**. **(B)** Muscle-IR is categorized based on Steady State Plasma Glucose (SSPG (mg/dL)) values (IS when SSPG≤120 and IR when SSPG>120). **(C)** β-cell function is categorized according to disposition index (DI, (pmol*dL)/(kg*ml)): normal when DI>2.2, intermediate when 1.2≤DI≤2.2, dysfunction when DI<1.2). **(D)** Incretin effect is categorized based on the measured incretin effect (IE) %: normal when IE>64, intermediate when 39≤IE≤64, and dysfunction when IE<39. **(E)** Hepatic-IR is categorized based on Hepatic-IR-index: IS when Hepatic-IR-index>3.95, intermediate when 3.95≤Hepatic-IR-index≤4.8, and IR when Hepatic-IR-index>4.8. **(F)** Heatmap showing the standardized deviance score of each metabolic measure for each participant (**Methods**). β-cell function and incretin effect values have been reversed so that higher positive values represent a greater abnormality. High positive deviance (darker red) indicates greater deviance from study population average in the abnormal direction (e.g., higher IR or lower incretin effect), while high negative deviance (lighter yellow) indicates a healthier metabolic phenotype than the average population. **(G)** Pairwise correlation network showing the association between the standardized deviance of the four metabolic measures. Pearson correlation coefficient (−1<r<1) was used to determine the strength of each relationship, where r=1 “dark orange” means strong positive correlation, and r=−1 “dark cyan’’ means strong negative correlation. The asterisk symbol on edges means that the correlation is statistically significant (p-value <0.05) between the corresponding two metabolic subphenotypes. **(H)** T2D polygenic risk score (PRS_T2D_) for all individuals in our cohort sorted in descending order and color by the glycemic status measured by HbA1c.

Insulin secretion rate was calculated using the widely used gold-standard C-peptide deconvolution method^41,42^, with C-peptide concentrations obtained at seven timepoints (0, 15, 30, 60, 90, 120, 180 min) during the OGTT. Insulin secretion was adjusted for insulin resistance (SSPG value) to generate a measure of β-cell function, referred to as disposition index (DI)^43,44^, also a validated gold-standard measure (see **Methods**). In this work, DI<1.58 (50th quartile in our cohort, **Supplementary Figure S1B**) indicates dysfunctional β-cell, whereas DI≥1.58 indicates normal β-cell function. **Figure 1C** demonstrates the heterogeneity of β-cell function (DI) in 8 participants at multiple time intervals during the OGTT (see **Supplementary Figure S2** for all participants, **Supplementary Table S2**). Participants S45 and S57 had a high DI (4.47 and 2.88, respectively) at early time intervals following the 75g oral glucose challenge, which demonstrates a “healthy” β-cell function. On the other hand, participants S28 and S58 had extremely low DI (0.67 and 0.98, respectively) following the oral glucose load, which indicates dysfunctional β-cells. As shown in Figure 2 A-C, β-cell function was generally higher among those with normoglycemia. Among those with prediabetes, the β-cell function varied widely.

The incretin effect (IE) was measured using the gold-standard isoglycemic intravenous glucose infusion test (IIGI)^45^ in which glucose is infused intravenously to mirror the glucose curve generated after oral glucose loading (see **Supplementary Figure S3** for concordance of glucose concentration between OGTT and IIGI). The difference in C-peptide concentrations during the OGTT relative to the IIGI was quantified at 7 time points (0, 15, 30, 60, 90, 120, and 180 min) to reflect the period of maximal hyperglycemia (**Supplementary Figure S4**). The degree to which C-peptide concentrations are elevated following oral versus intravenous glucose loading reflects the incretin effect (see **Methods, Supplementary Table S2**). In this work, IE<53.38% (50th quartile in our cohort, **Supplementary Figure S1C**) indicates dysfunctional incretin effect, whereas IE≥53.38 indicates normal incretin effect. The incretin response was extremely heterogeneous between participants and did not correlate with β-cell function nor insulin resistance as shown in **Figure 2D** and **2G**. **Figure 1D** depicts the C-peptide concentrations during oral (OGTT) as compared to intravenous (IIGI) glucose loading in six participants. Participants S42 and S32 had little difference in C-peptide responses during the OGTT and IIGI tests, indicating a poor incretin effect (IE of 4.80% and 23.28%, respectively). In contrast, participants S01 and S15 demonstrated a robust increase in C-peptide during OGTT versus IIGI, consistent with a high incretin effect (IE of 64.92% and 78.59%, respectively). Notably, the secretion of incretin hormones, as measured by glucagon-like peptide-1 (GLP-1) and gastric inhibitory polypeptide (GIP) concentrations, also varied considerably between participants (**Supplementary Figures S5A and S5B, respectively**). Interestingly, IE is significantly correlated with GIP at OGTT-2h GIP concentration (r=0.53, p-value=0.002), unlike GLP1 that is not significantly correlated with IE (r=0.3, p-value=0.1).

Hepatic insulin resistance (Hepatic IR) was calculated using the hepatic IR index, which is a formula validated against endogenous glucose production measured during euglycemic–hyperinsulinemic clamp^46,47^ (**Figure 2E, Methods, Supplementary Table S2**). In this work, hepatic IR index <4.35 (50th quartile in our cohort) indicates hepatic insulin sensitivity, whereas hepatic IR index≥4.35indicates hepatic IR (**Supplementary Figure S1D**). In our cohort, participants S58 and S44 had the highest hepatic IR with a hepatic IR index of 5.03 and 4.98, respectively. In contrast, participants S19 and S14 had the lowest hepatic IR with hepatic IR of 3.57 and 3.61, respectively.

### Dominant metabolic subphenotype calculation

For each participant, we attempted to determine which metabolic process was most impaired and thus classify individuals according to their “dominant” or codominant metabolic subphenotype(s). This classification would be useful clinically to inform order of treatment with different therapeutics that target disparate physiologic processes. Classification entailed calculating the standard deviation values of each of the four metabolic measures as the deviation from the cohort mean (**Methods**). Positive deviations from the mean (shown in red or dark red on the heat map in Figure 2F) indicate an unhealthy direction and negative deviations (shown as light orange or yellow on the heat map) indicate a healthy direction. **Figure 2F** highlights the differential deviance of each metabolic measure quantified per individual in our cohort. For each participant, the metabolic measure with the highest deviance (darker red) is the metabolic subphenotype that is more adversely affected relative to other participants. For example, for participant S42, the incretin effect was the metabolic subphenotype that was most negatively affected, and for participant S23, β-cell function was the most negatively affected - this method allows individuals to be classified according the measure with the greatest deviance, which was termed their metabolic subphenotype. The majority of individuals (n=16) had a single metabolic subphenotype that was most deviant (**Methods, Supplementary Table S3**), with β-cell function being most frequent (n=5), followed by muscle IR and incretin deficiency (n=4 each), and then hepatic IR (n=3). Thirteen participants had co-dominant metabolic subphenotypes (**Supplementary Table S3**), with hepatic IR and muscle IR, and β-cell and incretin deficiency having the highest prevalence (n=4 each). Only three individuals (S35, S47, S57) could not be classified by this method as having a dominant or co-dominant metabolic phenotype. At the cohort level, we observed that individuals with high levels of muscle IR also had high levels of hepatic IR (r=0.73, p-value=10^-5^) (**Figure 2G**). Indeed, a total of 13 individuals (34%) had either muscle, hepatic, or combined IR. A total of 13 individuals had dominance or co-dominance in β-cell dysfunction or incretin deficiency (40%). β-cell dysfunction was significantly correlated with muscle IR (r=0.6, p-value=2×10^-4^) but not with impaired incretin effect (r=0.33, p-value=0.06) (Figure 2G). Impaired incretin effect was not significantly associated with muscle or hepatic IR (r=0.25, p-value=0.17 and r=0.21, p-value=0.24, respectively). These data suggest that nearly all individuals in the normoglycemic or prediabetic state can be classified according to underlying metabolic physiology with similar balance across IR subphenotypes and insulin-secretion subphenotypes.

### Polygenic risk score as a predictor of glycemic level

Each participant was genotyped using a genome-wide array and the genetic predisposition for T2D was quantified by calculating the T2D polygenic risk score (PRS_T2D_) based on 338 independent signals for T2D-risk, as previously developed and validated ^49^ (**Methods, Supplementary Table S4**). **Figure 2H** shows that the PRS_T2D_ is associated with HbA1c level (r=0.48, p-value=0.005). A notable exception is female participant S07 (age at the time of the study 37 yrs, BMI 26.3kg/m^2^) who had a high PRS_T2D_ of 0.9, despite normoglycemic HbA1c (5.1%), muscle insulin sensitivity and β-cell function (SSPG=60 mg/dL, DI=2.58, **Figure 2B** and **2C**, respectively), and intermediate incretin effect and hepatic IR (IE=63.7%, hepatic IR index=4.3, **Figure 2D** and **2E**, respectively). As expected, PRS_T2D_ was positively correlated but did not reach a significance level, with SSPG (r=0.26, p-value=0.14), hepatic IR (r=0.28, p-value=0.1), HOMA-IR (r=0.3, p-value=0.08), BMI (r=0.18, p-value=0.29), and triglycerides (r=0.22, p-value=0.21). In addition, PRST2D was inversely correlated but did not reach significant levels, with DI (r=-0.2, p-value=0.25), IE (r=-0.16, p-value=0.36), HOMA-B (r=-0.32, p-value=0.06), and HDL (r=-0.28, p-value=0.12).

### Features of the glucose response curve during OGTT identify metabolic subphenotypes

Our main objective was to determine if we could predict metabolic subphenotypes from the glucose time-series response obtained from the standardized OGTT test (Supplementary Table S5). To achieve this we developed a robust machine learning framework in which features were extracted from the OGTT glucose time-series using two approaches: (a) engineered features from the OGTT glucose time-series (OGTT_G_Features) (b) a reduced representation method (OGTT_G_ReducedRep) that represents each normalized and smoothed OGTT curve with a vector of 2 dimensions (Figure 3A). In the first approach (OGTT_G_Features), we extracted 14 features from the OGTT glucose time-series (Methods). Figure 3B shows that multiple extracted features from OGTT glucose time-series were significantly correlated with metabolic subphenotypes (p-value<0.05, marked as x in Figure 3B). Area-Under-the-Curve (AUC) and incremental Area-Under-the-Curve (iAUC) were the two highest significantly positively correlated features with muscle IR (r=0.59 and p-value=3×10^-4^, r=0.57 and p-value=6×10^-4^, respectively). Glucose levels at 120 minutes (G_120_) and iAUC were the top two negatively correlated with β-cell function (r=-0.59 and p-value=4×10^-4^, r=-0.55 and p-value=3×10^-3^, respectively). Positive Area-Under-the-Curve (pAUC) and Glucose Peak levels (G_Peak) were the top two features significantly negatively correlated with the incretin effect (r=-0.72 and p-value=6×10^-5^, r=-0.68 and p-value=2×10^-5^, respectively). Finally, the slope from peak to the end at 180 min (S_peak2end) and baseline glucose levels (G_0_) were the top two parameters significantly correlated with the hepatic IR (r=-0.51 and p-value=0.002, r=0.47 and p-value=0.006, respectively).

**Figure 3.**
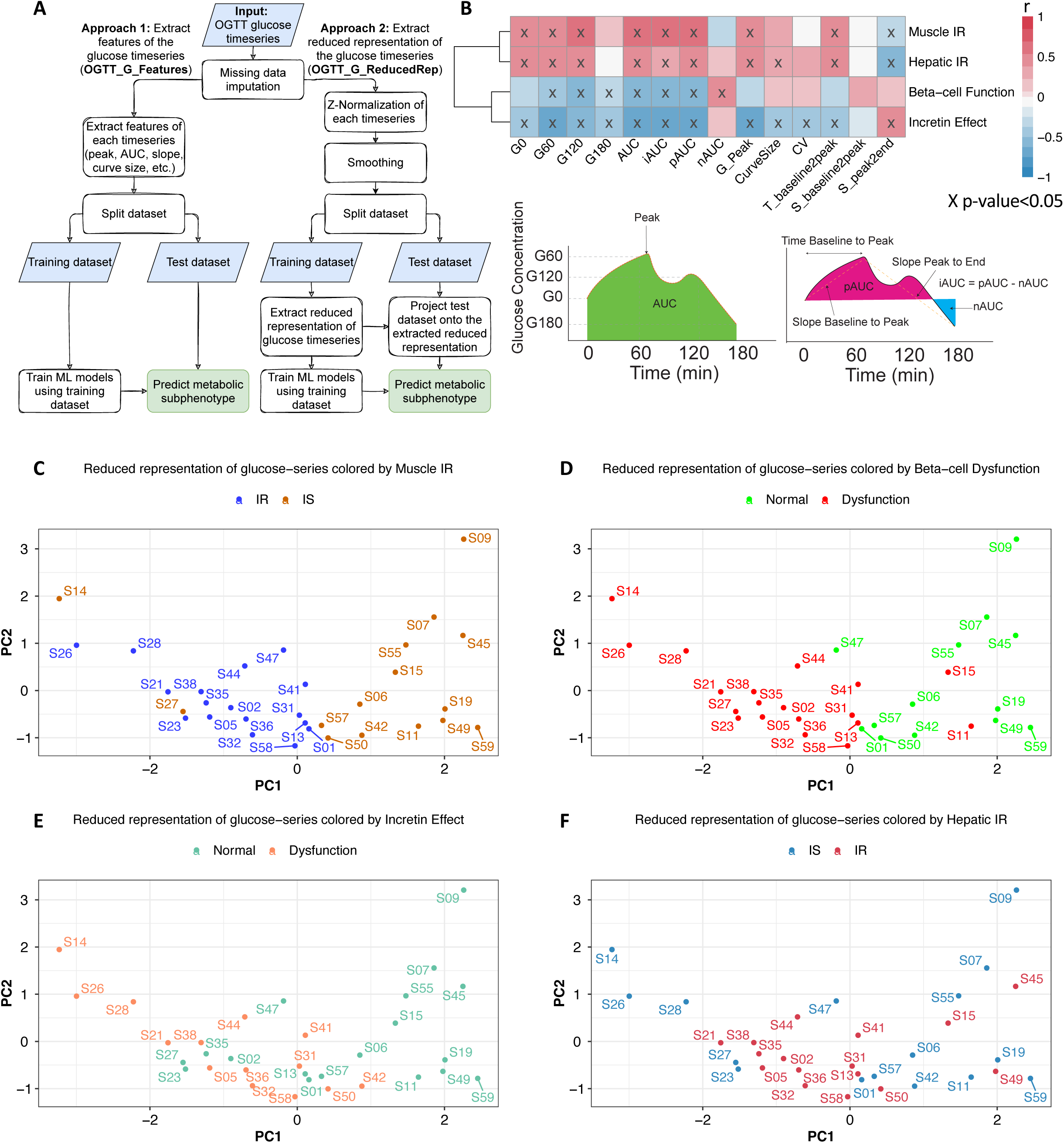
Features of the glucose time-series identify metabolic subphenotypes. **(A)** Machine learning framework for predicting metabolic subphenotype using OGTT glucose time-series. **(B)** Relationship between OGTT glucose time-series features and metabolic subphenotypes. Pearson correlation coefficient (−1<r<1) was used to determine the strength of each relationship, where r=1 “dark red” means strong positive correlation, and r=-1 “dark blue” means strong negative correlation. The “x” in some of the heatmap cells means that the correlation is statistically significant (p-value <0.05) between the curve feature and the corresponding metabolic subphenotype. At the bottom of the figure there is an illustration of some of the glucose curve features. *G_t_* denotes plasma glucose level at time *t,* AUC denotes area under the curve, iAUC denotes incremental area under the curve, pAUC denotes positive area under the curve, nAUC denotes negative area under the curve, and CV denotes coefficient of variation, peak glucose level (G_Peak), length of the glucose time-series (CurveSize), time from baseline to peak value (T_baseline2peak), slope between baseline to the peak glucose level (S_baseline2peak), and slope between glucose values at the peak and at the end (at t=180 min) (S_peak2end) (**Methods**). PCA of OGTT glucose time-series is colored according to classification using gold standard metabolic tests for **(C)** muscle IR (measured by SSPG), **(D)** β-cell function (measured by disposition index), **(E)** incretin effect, and **(F)** hepatic IR (inferred by hepatic IR index). The text on the plot represents the participant ID.

In the second approach (OGTT_G_ReducedRep), we first Z-normalized and smoothed the OGTT glucose time-series of 16 timepoints (Supplementary Figure S6). We then extracted the reduced representation of the OGTT glucose time-series (Methods). Interestingly, using the reduced representation of the OGTT glucose time-series, there was a very clear separation of muscle IR and IS (Figure 3C). Additionally, the reduced representation of OGTT glucose time-series readily distinguished the three classes of β-cell function (normal, intermediate, and dysfunction) (Figure 3D). Individuals with normal β-cell function were located on the positive side of the first principal component (PC1). In contrast, individuals with β-cell dysfunction were found on the negative side of PC1, and individuals with intermediate β-cell function are found in between. The same pattern was observed for incretin effect classes (Figure 3E), where participants with normal versus impaired incretin effect were reasonably well separated. Participants with an intermediate incretin effect overlapped with the two more extreme groups. On the other hand, hepatic IR classes were not separable using the reduced representation of the processed OGTT glucose time-series (Figure 3F).

We calculated the correlations between the top 2 PCs, OGTT_G_ReducedRep, and the extracted features, OGTT_G_Features (Supplementary Figure S7). We found a relationship between the first two PCs and the 14 extracted curve features. The first 2 PCs are used to reduce the dimension in feature space while capturing the major variances (43.8% and 18% for PC1 and PC2, respectively). For example, higher positive values on the PC1 are associated with lower iAUC, which from Figure 3B, lower iAUC was associated with muscle IS. Figure 4C also confirms this relationship whereby muscle IS participants are clustered towards higher positive values of PC1.

**Figure 4.**
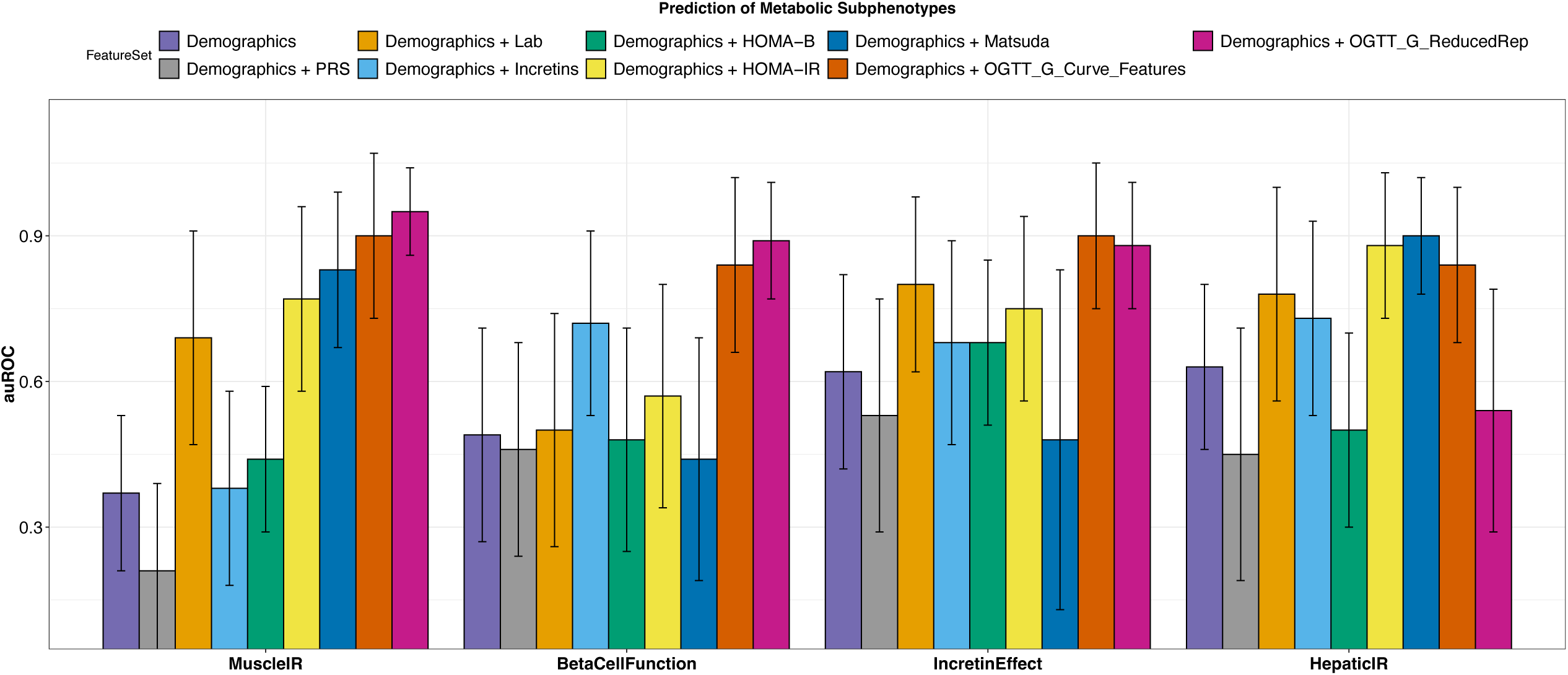
Benchmarking of metabolic subphenotyping prediction from features extracted from OGTT glucose time-series versus existing surrogate markers. Bar graphs represent the average area under ROC (auROC) of the best performing model for each metabolic subphenotype and each corresponding feature set. Error bars represent the standard deviation of the measured auROC. In total, 9 feature sets were evaluated for each metabolic subphenotype; two sets of features were obtained from OGTT glucose curve (OGTT_G_Features and OGTT_G_ReducedRep), T2D polygenic risk score (PRS), and six measures in current use including Demographics (age, sex, BMI, ethnicity, and participant family history for T2D), Demographics+Lab (demographic variables plus HbA1C, and FPG), HOMA-B (a surrogate marker for β-cell function), HOMA-IR and Matsuda Index (both are surrogate markers for muscle insulin resistance), and Incretins (total GIP and GLP-1 concentrations at OGTT_2h, which are optimized surrogate marker for incretin effect). Four classifiers were trained on the training set and the y-axis represents the auROC of the best performing classifier on the test set, for each metabolic subphenotype and each feature set. Statistical significance is performed between the measure of auROCs among all tested features and OGTT_G_ReducedRep using the Wilcoxon Rank Sum Test.

**Figure 5:**
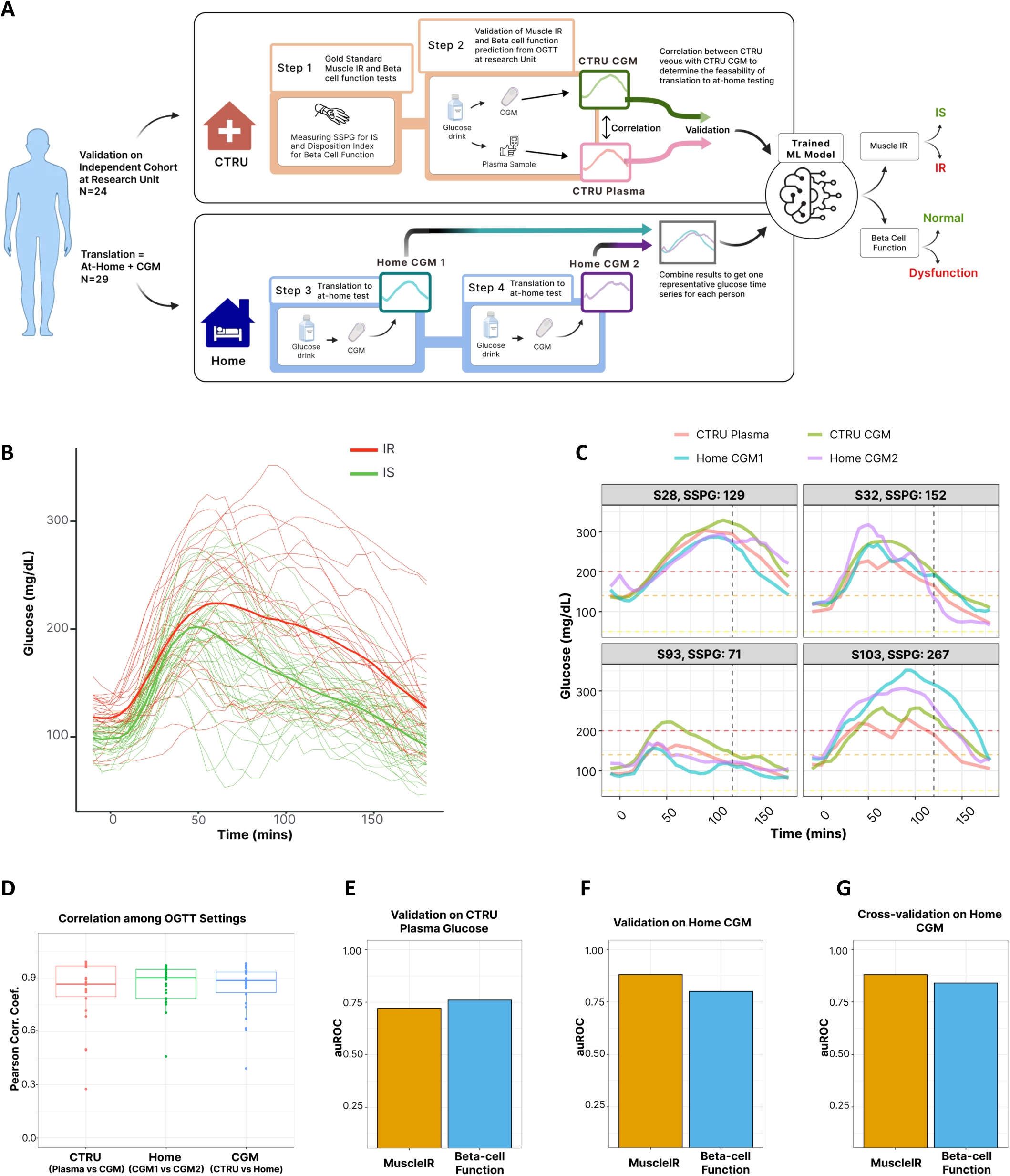
Validation on independent cohort and translational to at-home CGM testing. **(A)** Study design of the validation cohort and at-home OGTT test via CGM to predict muscle IR and β-cell function. **(B)** CGM glucose curves between IR (red) and IS (green) during an OGTT. Each group (IR or IS) CGM curve is visualized with the thicker line representing the mean curve. **(C)** Examples of four participants, each with four glucose curves; two curves from OGTT at clinical setting via venous sampling and CGM, and two curves from at-home OGTT via CGM. Subjects S28 and 93 are IS (SSPG<120 mg/dL), and subjects S32 and S103 are insulin resistant (SSPG>120 mg/dL) **(D)** Correlations among glucose time-series measured via different OGTT settings (clinical settings vs at-home, plasma vs interstitial via CGM, and reproducibility of 2 at home OGTT via CGM). Each point represents the correlation between two glucose series of the same person. High positive correlations indicate that time-series patterns are preserved among test settings. **(E)** Muscle IR and β-cell function prediction performance using plasma glucose series from the independent validation cohort during an OGTT at CTRU. **(F)** Muscle IR and β-cell function prediction performance on CGM glucose series from the independent validation cohort during an OGTT at Home. **(G)** Muscle IR and β-cell function prediction performance using cross-validation on CGM glucose series from the at-home cohort during an OGTT at Home.

### Predicting metabolic subphenotypes from glucose time-series using machine learning

The separability in the classes of muscle IR, β-cell function, and incretin effect, using the reduced representation of the processed OGTT glucose time-series suggested that a robust prediction of such classes could be achieved using a learning framework. For each feature extraction approach, OGTT_G_Features and OGTT_G_ReducedRep, the dataset was split into a training set and a test set. For each metabolic subphenotype, we optimized the hyperparameters of four different classifiers: Random Forest (RF), support vector machine with a radial basis function kernel (SVM-RBC), support vector machine with a linear kernel (SVM-linear), and logistic regression with L1 regularization (LR-L1). Models were evaluated comprehensively to ensure the generalizability and robustness of the trained models (**Methods**). We benchmarked the prediction of metabolic subphenotypes using the two sets of extracted features from the OGTT time-series by comparing performance to surrogate metabolic measures in current use including (1) Demographics (age, sex, BMI, ethnicity, and participant family history for T2D); (2) T2D PRS and Demographics; (3) Lab (HbA1c and FPG) and Demographics; (4) HOMA-B (**Supplementary Figure S8A**), a surrogate marker for β-cell function, and Demographics; (5) HOMA-IR (**Supplementary Figure S8B**), and Demographics; (6) Matsuda Index **(Supplementary Figure S8C**), surrogate markers for muscle IR; and Demographics (7) total GIP and GLP-1 concentrations at OGTT_2h (an optimized surrogate marker for incretin effect) and Demographics. The performance of each model, on each feature set, and each metabolic subphenotype was evaluated using the area under the receiver operating characteristic curve (auROC), sensitivity, specificity, precision, F1-score, and accuracy (**Supplementary Table S6**). For each metabolic subphenotype and for each feature set, models were ordered based on F1-score and then sensitivity, and the top model was selected as top performer. **Table 2** summarizes all performance metrics of the top performing model for each metabolic subphenotype, and for each tested feature set. Statistical significance was performed between the measure of auROCs among all tested features, and OGTT_G_Features and OGTT_G_ReducedRep using the Wilcoxon rank-sum test (**Supplementary Table S8**). All p-values were adjusted for multiple testing using Bonferroni method.

Figure 4 summarizes the auROC of the best performer classifier for metabolic subphenotype using each feature set. Muscle IR was predicted with extraordinarily high auROC=0.95 using OGTT_G_ReducedRep, while OGTT_G_Feature predicted Muscle IR with auROC=0.90. Importantly, OGTT_G_ReducedRep had significantly higher predictive power than any one of the currently used features to predict Muscle IR, including our optimized measure of Demographics+Lab (auROC=0.69, p-value=1.2×10^-19^), Demographics+HOMA-IR (auROC=0.77, p-value=7×10^-15^), and Demographics+Matsuda index (auROC=0.83, p-value=2.9×10^-8^). β-cell dysfunction was predicted from OGTT_G_ReducedRep with an auROC of 0.89, which is significantly higher than Demographics+Lab (auROC=0.5, p-value=7.6×10^-24^), HOMA-B (auROC=0.48, p-value=1.28×10^-25^), and Demographics+HOMA-IR (auROC=0.57, p-value=7.8×10^-20^). Incretin deficiency could be predicted with an auROC of 0.88 using OGTT_G_ReducedRep and auROC of 0.90 using the OGTT_G_Features, which was higher than Demographics+Incretins (auROC=0.68, p-value=2.6×10^-12^), and Demographics+Lab (auROC=0.8, p-vlaue=2.9×10^-3^). For Hepatic IR, OGTT_G_Features showed a promising performance (auROC=0.84), however Matusda Index (auROC= 0.90, p-value=0.86) and HOMA-IR (auROC=0.88, p-value=1) showed superior performance, but not statistical significance. Thus, models for different metabolic subphenotypes could be built from features of the OGTT glucose timeseries.

### Validation of metabolic subphenotype predictions in independent cohort

We next validated the prediction of muscle IR and β-cell function in a separate independent cohort of 24 individuals matched for age, sex, BMI, and HbA1c (Figure 5A, **Table 1, and Methods**). Frequently sampled OGTT as well as IST were conducted under standardized conditions in the CTRU as previously described to obtain gold standard measures of Muscle IR and β-cell function. The model focused on features extracted from glucose time series (OGTT_G_Features) as previously described and demonstrated in Figure 3A. For each participant, muscle IR and β-cell function were predicted using the best performing trained model from the initial cohort (Figure 4, **Methods**). On the independent validation cohort and using plasma glucose series obtained through OGTT at CTRU, muscle IR was predicted with auROC of 0.72, and β-cell function with auROC of 0.76 (Figure 5E).

### At-home prediction of muscle insulin resistance and β-cell function through the use of continuous glucose monitoring

To demonstrate the practical value of performing this test at home as well as the reproducibility of our proposed framework we sought to investigate the feasibility of utilizing at-home testing with CGM to predict muscle IR and β-cell function. Cohort characteristics of 29 individuals who were recruited from the main (n=5) and validation (n=24) cohorts are summarized in **Table 1**. Participants underwent gold-standard testing for insulin resistance (SSPG test) and B-cell function (16-point OGTT with c-peptide deconvolution adjusted for SSPG and expressed as DI) as described, as well as two OGTTs administered at home under standardized conditions during which glucose patterns were captured by a CGM within a single 10-day session (Dexcom G6 pro) (Figure 5A**, Supplementary Table S5**).

Participants consumed a 75g glucola (NERL Trutol, Thermo Fisher Scientific) in a 10 oz serving, (identical to method used in the CTRU OGTT) after an overnight fast of 10-12 hours. They were instructed to follow dietary and physical activity guidelines to standardize the test to minimize environmental influences on glucose response (**Methods**). CGM curves for participants defined by SSPG as IR or IS are shown in (Figure 5B). Even with visual inspection of the curves, clear differences between IR and IS individuals are apparent: as compared to IS, IR individuals had higher glucose peak (p<0.05), a broader glucose curve that remained high, and thus higher AUC. This pattern is consistent with our previous findings (Figure 3B) when plasma glucose levels were measured at various intervals in the CTRU.

We conducted a thorough investigation into the congruity of glucose time-series acquired through different methods of OGTT, including a) measurements of glucose levels from plasma samples collected after glucola consumption while simultaneously wearing a CGM, and b) in the home setting using CGM. Figure 5C displays a representative example of the glucose time-series for four participants, each of whom had four glucose time-series taken during different OGTT settings: two simultaneous series in the clinical research unit one measured venously and the other via CGM, and two series acquired via two repeated at-home OGTT using CGM (see **Supplementary Figure S9** for all time-series of all participants). Despite minor variations among the time-series, the pattern of the glucose time-series remained consistent across all settings. The concordance between different settings was quantified and depicted for each participant in Figure 5D. In general, the Pearson correlation between the glucose time-series obtained in the clinical setting through plasma and CGM measurements was 0.81 (p-value < 2.2×10^-16^); the correlation between the two at-home OGTT using CGM was 0.86 (p-value < 2.2×10^-16^); and the correlation between the clinical setting time-series measured venously and the mean of the two at-home OGTT time-series was 0.80 (p-value < 2.2×10^-16^). These high correlations indicate that CGM measurements performed at home are similar to those obtained in the clinical research unit and have high concordance with each other when performed under standardized conditions to minimize environmental perturbations.

Features from the glucose time series curves were extracted as previously described in Figure 3A. The classification of muscle IR (auROC=0.88) and β-cell function (auROC=0.80) based on the mean of two home CGM curves performed even better than plasma curves in the independent test set (Figure 5F). This improved performance is likely due to the efficient information extraction from both the feature engineering and the continuous measurements of CGM, which contains much more information than those from the venous sampling in which measurements are less frequent. A cross-validation based on CGM data alone showed similar performance for muscle IR (auROC=0.88), whereas the prediction of β-cell function was further improved (auROC=0.84) (Figure 5G).

## Discussion

T2D and its precursor, prediabetes, are currently defined on the basis of glycemic level rather than underlying pathophysiology. The mechanistic basis, however, is complex, and several distinct pathophysiological processes contribute. Here we demonstrate that individuals with glucose ranging from normoglycemic to prediabetes exhibit clear heterogeneity in four distinct physiologic processes that contribute to disordered glucose metabolism, including muscle IR, β-cell dysfunction, impaired incretin effect, and hepatic IR. We clearly showed that these phenotypes do not necessarily correlate with traditional glucose measures such as HbA1c and thus cannot be estimated in this way. Using gold-standard physiologic tests and expressing values for each individual as the relative deviance from the cohort mean, we expressed the degree to which each process was abnormal. The majority of individuals exhibited a single dominant metabolic subphenotype or two codominant phenotypes. These partitioned generally into IR dominant subphenotypes (muscle and liver IR), comprising 34% of the cohort, or insulin secretion subphenotypes (B-cell and incretin deficient), comprising 40% of the cohort. Muscle and hepatic IR, were highly intercorrelated and frequently exhibited co-dominance. This suggests that from a clinical classification perspective muscle and hepatic IR might be combined into one group classified as “insulin resistant”, and therapies known to improve insulin resistance (eg. dietary or pharmacologic weight loss (including GLP-1 or dual GLP-1/GIP agonists), physical activity, metformin, thiazolidinediones) used as initial treatment. β-cell and incretin deficiency phenotypes, together accounted for the majority of patients, might be amenable to similar treatment strategies that target insulin secretion, since this is the common denominator in the pathophysiology for both of these subphenotypes. These include agents that augment or replace insulin secretion or incretin action (e.g., sulfonylureas, glinides, DPP4 inhibitors, GLP-1 agonists or GLP1-GIP dual agonists, or insulin). Only 9% of participants could not be classified as having a dominant or co-dominant metabolic subphenotype, thus indicating that use of the glucose “shape of the curve” method to identify underlying physiology could inform targeted approaches for diabetes prevention or treatment.

Historically, subclassification of T2D into more specific subtypes has been limited by the lack of feasible scalable standardized diagnostic tests. The requirement for laborious and invasive research tests to characterize underlying metabolism precludes their use outside of a research setting and surrogates such as HOMA-IR only explain 40% of insulin resistance^40^ and are limited by lack of standardization of insulin concentrations across laboratories. Recent advances in understanding the underlying biology of glycemic disorders and the development of pharmacologic therapies targeting multiple distinct pathways to hyperglycemia renders a physiologically-based subclassification of prediabetes and T2D more important than ever. In fact, the scientific discovery regarding the “incretin” effect has led to the development of an entire new class of effective pharmacotherapies for T2D. Other classes of glucose lowering therapeutics, as well as dietary weight loss and exercise target insulin resistance, β-cell function, hepatic glucose production, and urinary glucose reabsorption. The majority of these medications and lifestyle interventions have been shown not only to reduce glucose in T2D, but also to significantly reduce risk of progression to T2D in individuals with prediabetes^14–16,48,49^. Thus, if pathophysiology could be identified in each individual, targeted therapy and a precision medicine approach would be possible. Given that 95% of the 13M individuals with diabetes are grouped into one category of T2D and the availability of an increasing array of therapeutic options, it is timely to consider subclassifying T2D. Prior attempts to do so based on simple laboratory tests or clinical/demographic markers that “cluster” together does not lend itself to a true precision approach to treatment however, which is based on targeting underlying physiology. Thus, the shape of the glucose curve using longitudinal measures and a machine learning approach represents the first successful subclassification according to underlying physiology and warrants testing in trials that evaluate the relative efficacy of targeted treatment. Our study is the first study to start with physiological subphenotypes and then seek a biomarker (glucose shape of the curve) that could be used in the clinic and at home.

A prerequisite for physiologic subclassification is a practical, accurate, accessible, and standardized test. Gold-standard tests for insulin resistance, insulin secretion, and incretin effect entail the use of intravenous infusions in the research unit are laborious, invasive and expensive and thus are not scalable nor practical. Hence, we seek to validate a more practical measure such as the shape of the glucose curve, particularly obtained by CGM, that could be used in the clinical setting. The OGTT test is relatively more feasible and has been widely studied with regard to its potential not only to identify diabetes based on a 2 hour glucose, but also to predict risk of developing T2D, CVD, and total mortality^29,30^. Furthermore, glucose obtained at two to five time points during OGTT has been correlated in multiple studies with underlying physiology^29,31–36^. Simple measures such as 1 hour glucose peak, time to peak, nadir below baseline, or relative height of 2 vs 1 hour or baseline glucose were noted to correlate with insulin resistance and β-cell function^31–33^. A number of publications showed that a mono versus biphasic curve shape was associated with insulin resistance and β-cell dysfunction and risk for diabetes^34–36^. Due to the simplicity of these analyses and the risk of missing the peak glucose with limited sampling and the high false positive rate given that the majority of normoglycemic individuals have a monophasic curve, more sophisticated modeling approaches have been employed^30,36,50^.

Functional principal components analysis analysis in pregnant women was able to differentially predict development of gestational diabetes^50^, and latent class trajectory analysis^36^ identified four curve types which differed according to degree of insulin resistance and β-cell function, and predicted diabetes, cardiovascular events and total mortality^30^. Results of the current study extend those of the prior studies by applying a machine learning framework to analyze the a dynamic glucose time series response to an OGTT with 16 time points, with demonstrated ability to identify insulin resistance, β-cell defect, and incretin defect better than currently-available tests including surrogate biomarkers, clinical-demographic profile, and polygenic risk score. This machine learning framework did not predict hepatic IR any better than HOMA-IR. However, given the high correlation between muscle and hepatic IR, we propose that it may be prudent to consider these two IR subphenotypes as a single IR phenotype. Despite the increasing potential of sophisticated approaches to leverage the OGTT to predict underlying metabolic phenotype, the OGTT test still poses a patient burden, limiting its widespread clinical use.

In the current study we aimed to evaluate whether a glucose curve generated by CGM would yield a similar ability to identify underlying metabolic physiology. Such test can be performed conveniently at home and is facile and inexpensive. Our results show that when applied to a CGM-generated glucose curve following standardized OGTT, the same machine-learning algorithm can accurately identify insulin resistance. The best performance was obtained by using the average of two home OGTTs performed in a single week under standardized conditions. Indeed, the average of two home CGM curves after OGTT performed better than plasma glucose values obtained following OGTT in the CTRU, presumably due to the high measurement frequency. Glucose curve features from at-home OGTT via CGM were extracted similarly to those from the frequently sampled OGTT in CTRU (Figure 3A). We have tested the Percent in Range (PIR) and Percent out of Range (POR) as possible features^51–54^, however, they did not increase the model performance. This is mainly because we used them during only a 180-minute session, and not the whole 10-day session. Also, PIR and POR are good for people with diabetes who usually have elevated glucose (hyperglycemia), or who are taking insulin and exhibit hypoglycemia. Our cohort is mainly normoglycemic and people with prediabetes. One major concern about the OGTT is the previously noted intra-individual variability^55^ of glucose at a given time point. This is in part likely but not completely due to environmental stressors (eg. eating a late dinner or snack, eating out, alcohol, late exercise or excessive duration of fasting, morning stress or coffee, change of location or time or rapidity of consumption of OGTT). We controlled for these with detailed patient instructions, and intrapatient variability was lowest for the home OGTT where the coefficient of variation was 11% for CGM across all time points. This compares favorably to the published 16.7% for 2-hour plasma glucose during OGTT^55^. The high reproducibility of home CGM may also also be due to similar conditions that can be maintained as well as the fact that we evaluated variability for the shape of the curve rather than a single time point, the latter of which may be more subject to variability as we noted that some curves were of similar shape but shifted up, down, right, or left compared to the other curve obtained from the same participant. If this method moves into the clinic for widespread use, it will be important to minimize environmental stressors that lead to variability in glycemic responses to the OGTT which can be accomplished with simple instructions for use.

Limitations of the current study include the relatively homogeneous demographics which are largely middle-aged Caucasian. Our BMI and glycemic range was sufficiently broad. Further studies to examine differences in predictive potential according to race, ethnicity and sex, as well as age and broader BMI subgroups would be informative. The current results do not extend to T2D beyond very mild cases with HbA1c of 6.5% or below (diet-controlled or new onset mild diabetes). Our two patients with diabetes were not previously diagnosed and had HbA1c of 6.5% and normal fasting glucose and thus are not reflective or more advanced diabetes. Additional studies will be required to determine if this method can be applied to individuals with T2D with more advanced hyperglycemia. At present, it seems the greatest utility of this method will be to diagnose underlying physiologic subphenotype in individuals at risk for diabetes. Also of note, we used a three-hour OGTT and the results may be different when compared to a two-hour OGTT. Secondly, while the reproducibility of the OGTT is historically imperfect, we showed high correlation between the two CGM curves generated during home OGTT, with an acceptable coefficient of variation (11%), likely due to our efforts to ensure standardization of environmental conditions. It will be important to have two of these done under standardized conditions and if one is vastly different from the first, a third should be done and the outlier excluded due to presumed environmental confounder. The optimal number of required at-home tests can be determined in future studies. Additionally, the hepatic IR index used in this study is based on a formula that was validated against a gold-standard test rather than being a gold-standard test itself <u>(Vangipurapu et al. 2011)</u>. Thus, it is less precise than our other metabolic measures, which are highly precise. This likely explains the suboptimal performance in predicting hepatic IR from glucose timeseries, compared to HOMA-IR or Matsuda index.

In summary, we demonstrate that the metabolic physiology underlying glycemic disorders varies widely between individuals and that dominant or co-dominant metabolic subphenotypes exist, which has implications for precision approaches to treat/prevent T2D given that interventions exist that target the separate physiologic components. We further show that modeling a glucose time-series response to oral glucose challenge can identify these metabolic subphenotypes. The features of the glucose time-series can identify muscle IR, β-cell deficiency, and incretin deficiency with superior predictive power than standard clinical and laboratory measures, accepted plasma surrogate markers, and polygenic risk score. We further show that prediction is even stronger using the average of two CGM glucose curves generated during home OGTT administered under controlled conditions to minimize environmental confounding. We thus suggest that identification of distinct metabolic subphenotypes using features of the glucose time-series during OGTT, including home OGTT with CGM, may enhance early identification of at-risk individuals who can then undergo targeted pharmacologic and lifestyle modification to prevent T2D. These results demonstrate how new technologies can unleash the potential of classic tests for metabolic health. If the patient burden posed by multiple blood draws over a 3 hour window is removed, the increased practicality and feasibility of OGTT at home may yield a metabolic test suitable for widespread use to identify metabolic phenotypes for risk stratification and targeted treatment. Thus, the current results provide support for the use of CGM to define metabolic subphenotypes based on the glucose shape of the curve during OGTT, and warrant trials to evaluate the relative efficacy of targeted treatment based on identified subphenotypes.

## Methods

### Study participants

The study protocol and clinical investigation were approved by the Stanford University School of Medicine Human Research Protection Office (Institutional Review Board # 43883). Participants provided written informed consent. Participants were recruited from the San Francisco Bay Area via locally placed advertisements online, in local newspapers, and during faculty lectures to the community. Participants were screened with a medical history and physical examination in the Stanford University Clinical and Translational Research Unit (CTRU), where vital signs, fasting plasma blood glucose, and baseline hematocrit, ALT, and creatinine were obtained. Eligibility criteria included age 30-70 years, BMI 23-40 kg/m^2^, absence of major organ disease, absence of type 2 diabetes defined by personal history or fasting glucose ≥ 126 mg/dL, uncontrolled hypertension, malignancy, chronic inflammatory conditions, use of any medications known to alter blood glucose or insulin sensitivity, hematocrit < 30, creatinine above the normal range, and ALT > two-fold above the upper limit of the normal range.

#### Initial cohort

Thirty-six participants enrolled and thirty-two completed all metabolic tests and were included in the final analyses as the main study cohort (also called training cohort) (**Table 1** and **Supplementary Table S1**).

#### Validation cohort

An independent cohort of 24 individuals completed OGTT and IST in the CTRU, as well as measuring HbA1c and lipid panel.

#### At-home CGM cohort

The validation cohort plus five of the initial cohort (n=29) was used for CGM analysis both in the CTRU (they were measured simultaneously during the OGTT test with plasma sampling and a blinded Dexcom G6 CGM). These individuals performed two at-home glucose tolerance tests on separate mornings under standardized conditions (see below).

### Quantitative metabolic physiological tests

All tests were performed in the Stanford CTRU after a ten-hour overnight fast.

#### A) Frequently-sampled oral glucose tolerance test (OGTT)

Participants were instructed to avoid celebratory or restrictive eating and unusual physical activity for 3 days prior to performing the test, fast 10-12 hours prior to the test, no strenuous exercise after 5 pm the evening before the test, and no smoking or using a nicotine patch the morning of the test. Dynamic plasma glucose profiles were obtained after administration of a 75t oral glucose load administered over 5 minutes (Brand Trutol, Manufacturer Fisher, 75g in a 10 oz serving). Plasma samples were drawn from an antecubital intravenous catheter drawn at 16 timepoints (−10, 0, 10, 15, 20, 30, 40, 50, 60, 75, 90, 105, 120, 135, 150, and 180 min). Glucose level was measured at the 16 timepoints (using the oximetric method), insulin and C-peptide (using the Millipore radioimmunoassay assay), both at 7 timepoints (0, 15, 30, 60, 90, 120, 180 min), GLP-1 (using Millipore ELISA EZGLP1T-36K kit), GIP (using Millipore ELISA EZHGIP-54K kit), Glucagon (using Mercodia ELISA 10-1271-01 kit), all at 4 timepoints (0, 30, 60, 120 min). Insulin, C-peptide, GLP-1, GIP, and Glucagon were measured at the Core Lab for Clinical Studies, Washington University School of Medicine in St. Louis. From the baseline sample (at 0 min), HbA1c, fasting plasma glucose, insulin, triglyceride, and high-density lipoprotein cholesterol levels were determined.

#### B) At home OGTT with CGM

Participants wearing CGM were instructed to do two at-home OGTTs that were standardized in order to minimize environmental influences that might lead to variability in results. The two at-home OGTT were performed within a single CGM sensor session (10 days) After an overnight fast, a 75 gram glucola drink (NERL Trutol, Manufacturer Thermo Fisher Scientific), was consumed in 10 oz over 5 minutes, and for three hours no oral intake other than water was allowed, and the patients were remained seated, identical to the OGTT procedure performed in the CTRU. Participants were required to do this test during a typical week that was devoid of celebratory or restrictive eating (e.g., wedding or religious holiday), with usual activity patterns (no unusual activity/training outside their typical patterns), and to refrain from eating out or performing strenuous exercise after 5 pm the night before the test, and not to eat or consume anything other than water after 10 pm on the evening before and morning prior to the test, and to avoid smoking or using a nicotine patch the morning of the test.

#### C) Isoglycemic intravenous glucose infusion (IIGI)

IIGI, as previously described<u>(Nauck et al. 1986)</u>, was performed in order to quantify the incretin effect by duplicating the plasma glucose profile during the corresponding OGTT. During the IIGI test, an intravenous catheter was placed in the antecubital vein for administration of continuous dextrose infusion at a rate needed to obtain desired glucose, which for each time point was the glucose level obtained at the same time point during the OGTT. Blood sampling from a second intravenous catheter included the same time points and assays as described above for the OGTT.

#### D) Insulin suppression test (IST)

Insulin-mediated glucose disposal was quantified by the modified insulin suppression test as originally described and validated^37–39^. Briefly, participants were infused for 240 min with octreotide (0.27 μg m^-2^ min^-1^) (to suppress endogenous insulin secretion), insulin (32 mU m^-2^ min^-1^), and glucose (267 mg m^2^ min^-1^). Blood was drawn at 30 min intervals for monitoring and at 10-min intervals from 210 to 240 min of the infusion to measure plasma glucose and insulin concentrations, and the mean of these four values was used as the steady-state plasma insulin (SSPI) and glucose (SSPG) concentrations for each individual. As SSPI concentrations are similar in all participants during these tests, the SSPG concentration provides a direct measure of the ability of insulin to mediate disposal of an infused glucose load; the higher the SSPG concentration, the more insulin-resistant the individual.

#### E) Measurement of plasma glucose

Blood was drawn from an intravenous catheter placed in the antecubital fossa which was warmed with a heating pad to obtain arterialized venous blood. The blood sample was run to the CTRU lab, spun for 2 minutes and plasma was analyzed on the YSI 2500 which is calibrated to the clinical laboratory glucose values at our institution. This method allowed for immediate bedside glucose results so that for the IIGI (below) the glucose infusion rate could be altered every 5 minutes as needed to mimic the glucose curve generated by the OGTT.

### Metabolic calculations

#### A) Muscle insulin resistance (SSPG)

While SSPG as described above reflects whole-body insulin-mediated glucose disposal, 85% of this occurs in muscle <u>(Reaven</u> <u>1988)</u>and thus, to distinguish this measure from hepatic insulin resistance, the term “muscle” insulin resistance is used in this study. Furthermore, a cutoff of < 120 mg/dL is generally indicative of insulin sensitivity. For the purpose of this paper, we defined non-insulin sensitive individuals (those with SSPG ≥ 120 mg/dL) as insulin resistance (IR). This includes individuals with moderate and severe IR, all of whom have been shown to respond to treatments known to improve insulin sensitivity (both lifestyle and medications).

#### B) β-cell function (Disposition Index [DI])

In order to quantify the status of the β-cell function, first calculated insulin secretion rate using C-peptide deconvolution method, using C-peptide concentrations measured during OGTT tests at 0, 15, and 30 mins. The Insulin SECretion (ISEC) software <u>(Hovorka et al. 1996)</u> was used to calculate prehepatic insulin secretion from plasma C-peptide measurements with adjustment for age, sex, and BMI <u>(Van Cauter et al. 1992)</u>. The ISEC software can be obtained from the author Roman Hovorka, PhD, Metabolic Modelling Group, Centre for Measurement and Information in Medicine, Department of System Science, City University, Northampton Square, London EC1V OHB, United Kingdom. We required an error coefficient of variation of 5% and 15-minute intervals. As the program uses a population model of C-peptide kinetics, we classified individuals as diabetic (“niddm”) if they had either fasting plasma glucose > 126 mg/dL, 2-hour glucose during OGTT > 200, or HbA1C > 6.5; or as “obese” if they were nondiabetic with BMI > 30. The remaining individuals were classified as “normal”. Because insulin secretion is best interpreted relative to insulin resistance (insulin resistant individuals need to secrete more insulin in order to maintain normoglycemia), we then calculated a disposition index (DI)^43,44^, as previously described, which adjusts the insulin secretion rate for degree of insulin resistance to generate an overall measure of B-cell function. DI is calculated as the area under the insulin secretion rate, divided by the SSPG (**Eq. 1**). DI thus describes β-cell function relative to insulin resistance.

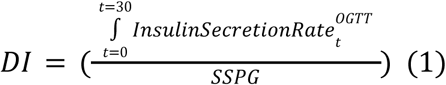

#### C) Incretin effect (IE%)

We quantified the incretin effect as previously described <u>(Nauck et al. 1986)</u> using the following (**Eq. 2**). Incretin effect is calculated based on the difference in the C-peptide concentrations during a frequently sampled OGTT and IIGI (0-180 min) that by design induce nearly identical plasma glucose concentrations. The difference in c-peptide in response to oral versus intravenous glucose loading reflects the contribution of incretin hormones from the gut that stimulate pancreatic β-cell insulin secretion. A higher incretin effect may indicate either increased secretion of incretin hormones (e.g., GLP-1 and GIP) and/or β-cell responsiveness to these hormones. Lower incretin effect may be due to deficiency in incretin hormones secretion and/or relative inability to stimulate insulin secretion from the β-cell.

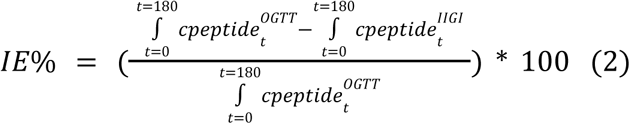

#### D) Hepatic IR index

Liver insulin resistance was inferred using a surrogate index as shown in **Eq. 3** <u>(Vangipurapu et al. 2011)</u>, where insulin is measured in (microIU/mL), plasma HDL cholesterol is measured in (mg/dL), and BMI is measured in kg/m^2^. In adults, Body Fat % (BF%) can be estimated using the Deurenberg Index as shown in **Eq. 4** ^56^, where BMI is measured in kg/m^2^, Age in years, and Sex = 1 when male, and Sex=0 when female.

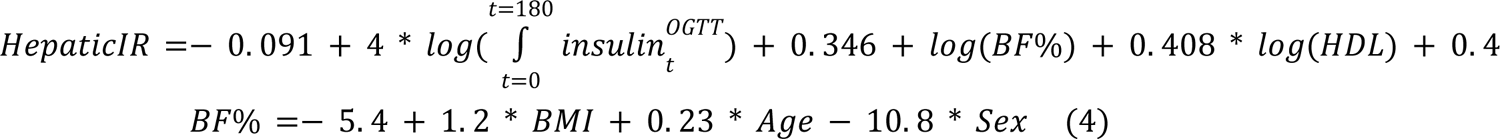

### Quantifying the relative deviance for each physiologic measure for metabolic subphenotype assignment

In order to classify each participant to their most “abnormal” physiologic process, we calculated a standardized deviance score (i.e. a Z-score) for each metabolic measure for each participant (**Eq. 5**), where msp^i^_j_ is the *ith* metabolic measure of participant *j* and mean(msp^i^) is the average of *i^th^* metabolic measure among the cohort’s participants and sd(msp^i^) is the standard deviation. High positive deviance indicates a more abnormal score, and stronger phenotype for that measure in a given individual; high negative deviance indicates a healthier metabolic measure, and thus relative absence of the unhealthy phenotype as compared to the average population. Disposition index (DI) and incretin effect (IE%) were negated before applying **Eq. 5**, since higher DI and IE% indicate healthier β-cell function and incretin function, respectively. As shown in the heatmap in Figure 2, it is possible to see in a given individual the relative deviance for each metabolic measure: some individuals had a single dominant (most deviant) measure, whereas others had two or more that were equally dominant.

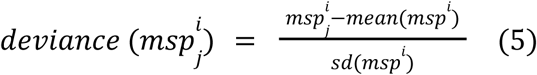

For each participant, the dominant metabolic subphenotype was identified based on the one exhibiting the highest deviance score. When the difference in deviance scores between the two highest metabolic subphenotypes was less than 0.5, the participant was classified as co-dominant. If more than one subphenotype exhibited deviance scores within 0.5 of the highest score, the participant was classified as neither dominant nor co-dominant. This method is not intended for the clinical diagnosis of metabolic abnormalities. Instead, it focuses on identifying individuals with deviations in metabolic profiles relative to a reference cohort. Further validation in diverse populations is required to assess its generalizability.

### Genotyping and T2D polygenic risk score calculation

DNA was extracted from whole blood peripheral blood mononuclear cells (PBMC). The concentration of the extracted DNA was measured using Qubit, normalized to 50 ng/uL, and 10 uL was used for each sample for genotyping. All samples were genotyped using the Illumina Infinium Omni2.5Exome array with ∼2.6 million single nucleotide variants (SNV) sites measured. Genotyping data underwent standard quality control using PLINK^57^, including filtering of SNVs with excess missing genotypes (>2%), filtering of SNVs out of Hardy-Weinberg equilibrium (p-value<1×10^-6^) and removal of samples with either excess missing genotypes (>3%) or discordant genetic vs recorded sex. Coordinates were aligned to the positive strand on genome build GRCh38 and the data was imputed against the TOPMed R2 reference panel^58–60^. After quality control and imputation, 11.8 million high-quality SNVs (R^2^>0.3) were obtained. T2D polygenic risk score (PRS_T2D_) was generated as an additive model (sum genotype dosage * weight) using 338 index variants and corresponding weights from the DIAMANTE multi-ancestry meta-analysis study^61^. Of the 338 variants, 16 were not available, for which mean population scores were added using the TOPMed Bravo platform reference frequencies (2*weight*population allele frequency)^60^.

### Machine learning framework to identity metabolic subphenotypes from OGTT glucose time-series

#### Preprocessing

Glucose time-series are preprocessed by imputing missing data using linear interpolation.

#### Features extraction

We extracted features for the machine learning framework using two feature extraction approaches (a) OGTT glucose time-series features (OGTT_G_Features) and (b) reduced representation of the OGTT glucose time-series (OGTT_G_ReducedRep). In the first approach (OGTT_G_Features), we extracted 14 features from the imputed OGTT glucose time-series, such as glucose level at time 0 (G_0_), 60 (G_60_), 120 (G_120_), 180 (G_180_), peak glucose level (G_Peak), length of the glucose time-series curve over the frequently sampled OGTT time interval (CurveSize), area under the curve (AUC), positive area under the curve (pAUC), negative area under the curve (nAUC), incremental area under the curve (iAUC), coefficient of variation (CV), time from baseline to peak value (T_baseline2peak), slope between baseline to the peak glucose level (S_baseline2peak), and slope between glucose values at the peak and at the end (at t=180 min) (S_peak2end). These 14 features demonstrated statistical significance in their Pearson correlation coefficients with one or more metabolic subphenotype indicators (SSPG, DI, IE, hepatic IR index). The only exception was “S_baseline2peak,” which exhibited a near-significant correlation with βeta-cell function (pvalue=0.06). In the second approach (OGTT_G_ReducedRep), we first Z-normalized the imputed OGTT glucose time-series to extract features related to curve patterns rather than the amplitude, then we smoothed the Z-normalized glucose time-series using a cubic smoothing spline with a smoothing parameter = 0.35. Then, the data was split into a training and testing set. For the testing set, we extracted the reduced representation of the OGTT glucose time-series using principal component analysis and the top two principal components were extracted to be used in training machine learning classifiers (OGTT_G_ReducedRep). For each new glucose series in the test set (*X*), the series is centered first using the mean vector of the training data (*X* _*centered*_). The dimension of the mean vector is 1×16, since we have 16 glucose timepoints per series. Afterwards, the centered series is multiplied by the loading matrix (*W*) of the top two principal components extracted from the training set (**Eq. 6**). The dimension of *W* is 16×2. *X*_*reduced*_ is the projected test series on the reduced representation space of the training set.

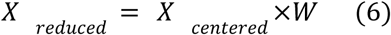

#### Training machine learning models

we next developed a machine learning approach, where the two extracted sets of glucose time-series features (OGTT_G_Features and OGTT_G_ReducedRep) were tested independently. For each feature extraction approach, the dataset was split into a training set and a test set. Data are stratified based on metabolic subphenotype class to ensure equal distribution of each class between training and test set. Participants from the initial venously sampled OGTT collection at CTRU without CGM data were assigned to the training set. No participants or any sample exists in both training or test sets. Cross validation is implemented on the training set.

For each metabolic subphenotype, we optimized the hyperparameters of 4 different classifiers: support vector machine with a radial basis function kernel (SVM-RBC), support vector machine with a linear kernel (SVM-linear), logistic regression with L1 regularization (LR-L1), and random forest (RF). We used the python package sklearn to train, tune, and test machine learning models. LR-L1 was trained using a SAGA solver and ran for a maximum of 10000 iterations to converge, with a tunable regularization strength hyperparameters C. SVM-RBF and SVM-Linear, were trained the sklearn.svm.SVC method, with kernel “linear” and “rbf”, respectively. The regularization parameter “C” is the tunable hyperparameter of both SVM models (linear and RBF). The tunable hyperparameters for random forest is the number of estimators. We have a grid of tunable hyperparameters (5 values) and search the best one based on cross-validation. All models are controlled by a preset random seed. The total number of models are 20 (4 basic model architectures and 5 hyperparameter values for each model).

Throughout the manuscript, all machine learning experiments were conducted using 5-fold cross-validation and are iterated 100 times. This entailed dividing the training set of plasma glucose from OGTT at CTRU (32 participants) into 5 groups, where 80% were used for model training, and 20% were used for validation during each fold. In each fold, we calculated the area under the receiver operating curve (auROC) together with other metrics. To ensure a more rigorous evaluation and stable model, we repeated the above cross-validation 100 times, where we shuffled the data before we divided the data into folds. This ensures that the performance of the framework is generalizable to any division of the data. Different hyperparameters are tested for each model in cross-validation and the order is shuffled after each iteration. The final model with selected hyperparameter value is chosen based on best auROC in cross-validation. The process of hyperparameter tuning and model selection is merged in the same loop because of the limited training sample size. However, the shuffle process ensures an accurate estimation of performance. The model is then tested on the independent test set on both data similar to the training set (plasma glucose from OGTT at CTRU) and CGM data. The class imbalance problem was handled via stratified sampling to ensure that training and validation contained the same proportion of class samples. Importantly, in the second approach of feature extraction (OGTT_G_ReducedRep), reduced representation was obtained from the training set, not the whole dataset, and used to train each classifier. The test set was then projected onto the reduced representation space obtained from the trained dataset, and their representations were used to predict metabolic subphenotypes. Obtaining reduced representation space from the training dataset instead of the whole dataset was crucial for the robustness and generalizability of the model by preventing data leakage between the training dataset and the test dataset.

We then conducted a comprehensive experiment to predict metabolic subphenotypes using the two sets of extracted features as previously discussed (OGTT_G_Features and OGTT_G_ReducedRep). For comparison, we evaluated the performance of these two feature sets to surrogate markers for metabolic phenotypes that are in current use, including (1) Demographics (age, sex, ethnicity, BMI, family history of T2D), (2) T2D Polygenic risk score, (3) Incretins (total GIP and GLP-1 concentration at hour 2 of the OGTT_2h), (4) Laboratory (HbA1C, and FPG), (5) HOMA-B^62^, (6) HOMA-IR^62^, and (7) Matsuda Index^63^. Demographics was evaluated alone and in all other models including our OGTT_G_Features and OGTT_G_ReducedRep models.

The performance of each model, on each feature set, and on each metabolic subphenotype was evaluated using the area under the receiver operating characteristic curve (auROC). We also calculated sensitivity, specificity, F1, precision, and accuracy. Metrics are aggregated and summarized.

#### Model validation on an independent cohort of OGTT at research unit

The independent validation cohort was first preprocessed then used to extract the two sets of glucose time series features similarly to the initial cohort. Then, trained models from the initial cohort were used to predict muscle IR.

### Statistical analyses

Statistical significance was performed using Wilcoxon rank-sum test with Bonferroni correction when multiple testing is encountered, such as the case with evaluating the metabolic subphenotypes predictions using different feature sets. All p-values mentioned in this work refer to p-values after Bonferroni correction.

#### Visualization

We used the R packages ggplot2 (v3.3.2) and pheatmap (v1.0.12) to plot most of the figures^51^. Summary Figure, Figure 1A, and Figure 5A were created using the Figma design tool. (https://www.figma.com/). Figure 3A was created using the draw.io platform (https://app.diagrams.net).

## Data and code availability

All data and code will be made publicly available upon manuscript acceptance. This study is part of the Precision Diets for Diabetes Prevention clinical trial (NCT03919877).

## Data Availability

All data and code will be made publicly available upon manuscript acceptance

## Acknowledgments

We thank Alessandra Breschi, Lettie McGuire, Susan Kirkpatrick, Monika Avina, Melanie Ashland, Kexin Cha, Jou-Ho Shih, Nicole Turk, and Varsha Rajesh for their valuable efforts throughout the project. This work was supported by NIH/NIDDK R01 DK110186-01 and a Stanford PHIND award. We acknowledge the support of the Stanford Diabetes Research Center (P30DK116074). ALG is funded by the Wellcome Trust (200837) and National Institute of Diabetes and Digestive and Kidney Diseases (NIDDK) (U01-DK105535, U01-DK085545, UM1DK126185). YW was supported by the American Diabetes Association Grant 11-23-PDF-76. AAM is currently an employee of Google. DP and TM are members of the scientific advisory board of January AI. ALG’s spouse is an employee of Genenetch and holds stock options in Roche. MPS is a co-founder and a member of the scientific advisory board of Personalis, Qbio, January AI, SensOmics, Protos, and Mirvie. He is on the scientific advisory board of Danaher, GenapSys, and Jupiter. No other authors have competing interests.

## Main Table Legends

**Table 1:** Baseline characteristics of the study cohorts. Continuous variables are reported as average ± standard deviation and categorical variables as frequency.

**Table 2:** Performance metrics of best models for each metabolic subphenotype, for each feature set.

## Supplementary Table Legends

**Table S1:** Participants’ Demographics.

**Table S2:** Initial Cohort Participants’ Calculated Metabolic Indicators.

**Table S3:** Deviance score (Z-score) and identification of dominant/Co-dominant metabolic subphenotypes.

**Table S4:** T2D Standardized Polygenic Risk Score (PRS) for the Initial Cohort.

**Table S5:** Glucose Timeseries Measurements for All Cohorts (CTRU and at-Home).

**Table S6:** The performance of predicting metabolic subphenotype using various feature sets and ML classifiers.

**Table S7:** The performance of predicting metabolic subphenotype using various feature sets and ML classifiers (without Demographics Combined into Glucose Features and Surrogate Biomarkers)

**Table S8:** Statistical significance analysis between auROCs among all tested features, and OGTT_G_Features and OGTT_G_ReducedRep using the Wilcoxon Rank-Sum Test. All p-values were adjusted for multiple testing using Bonferroni method.

## Supplementary Figures Legends

**Figure S1:**
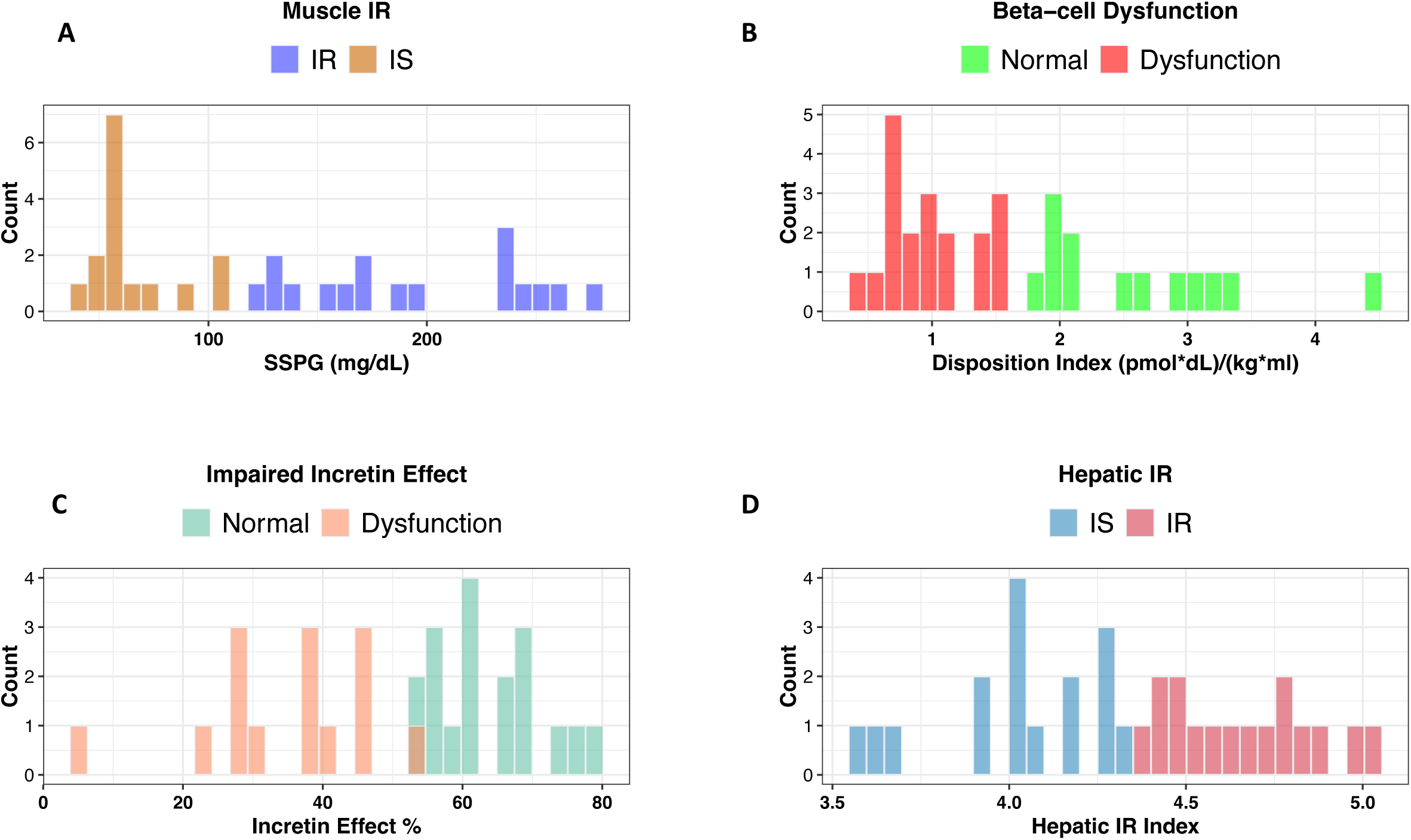
Distributions of classes of each metabolic subphenotype in the initial cohort. **(A)** muscle insulin resistance is categorized based on steady-state plasma glucose (SSPG), **(B)** β-cell dysfunction is categorized based on disposition index (DI), **(C)** impaired incretin effect is categorized based on the area under the C-peptide curves during OGTT, and IIGI, and **(D)** hepatic insulin resistance is categorized based on the hepatic insulin resistance index.

**Figure S2:**
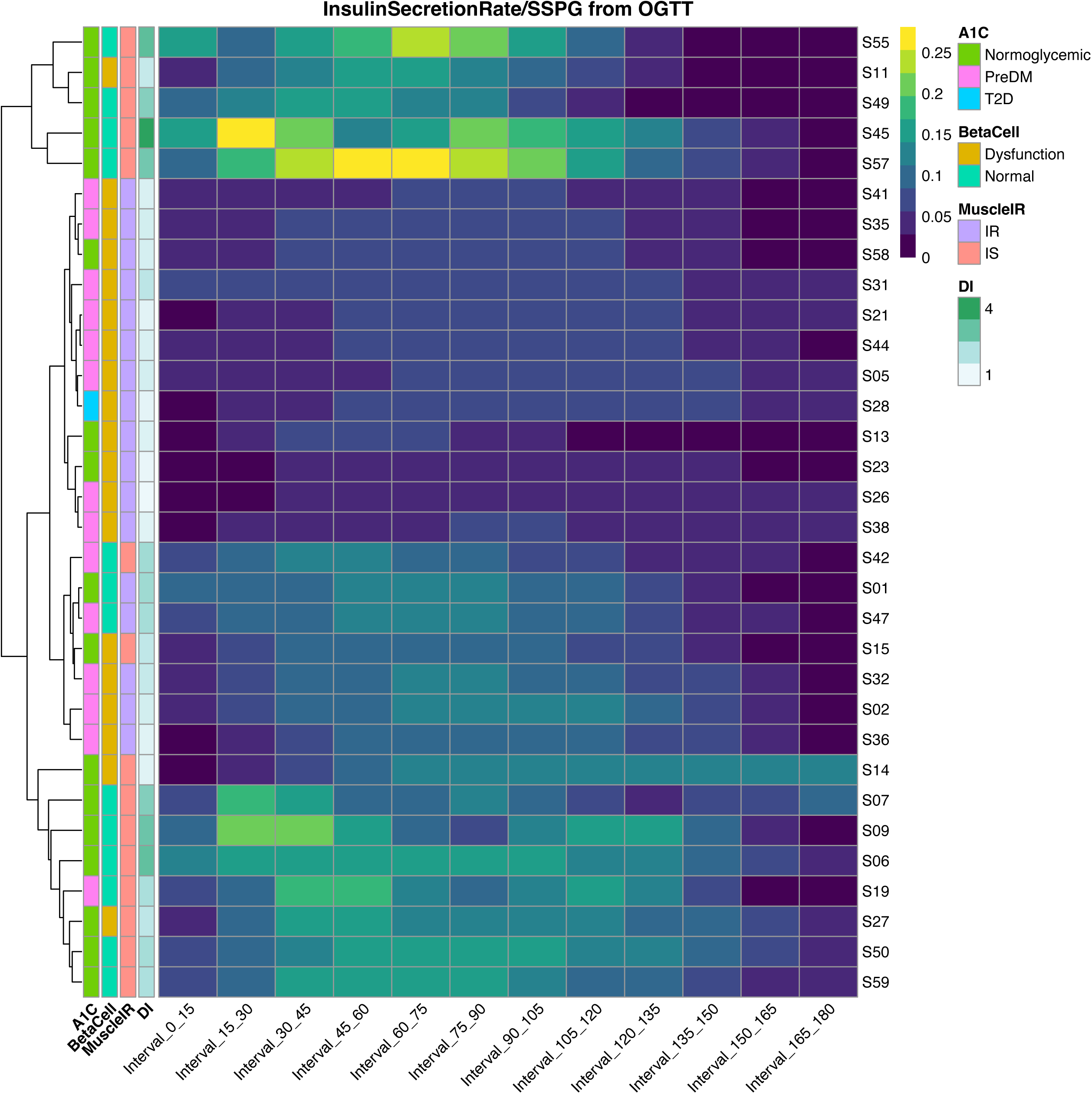
Insulin secretion rate measured during OGTT normalized by SSPG used to infer β-cell function. The insulin secretion rate was calculated using the C-peptide deconvolution method with C-peptide concentrations obtained at seven timepoints (0, 15, 30, 60, 90, 120, 180 min) during the OGTT, which was then adjusted for insulin resistance to generate a measure of β-cell function. Rows represent each participant, and columns represent time intervals in minutes relative to the start of OGTT. Yellow cells indicate time intervals with higher insulin secretion. Each participant is annotated with HbA1C class, β-cell function class (BetaCell), muscle insulin resistance class (MuscleIR), and disposition index (DI).

**Figure S3:**
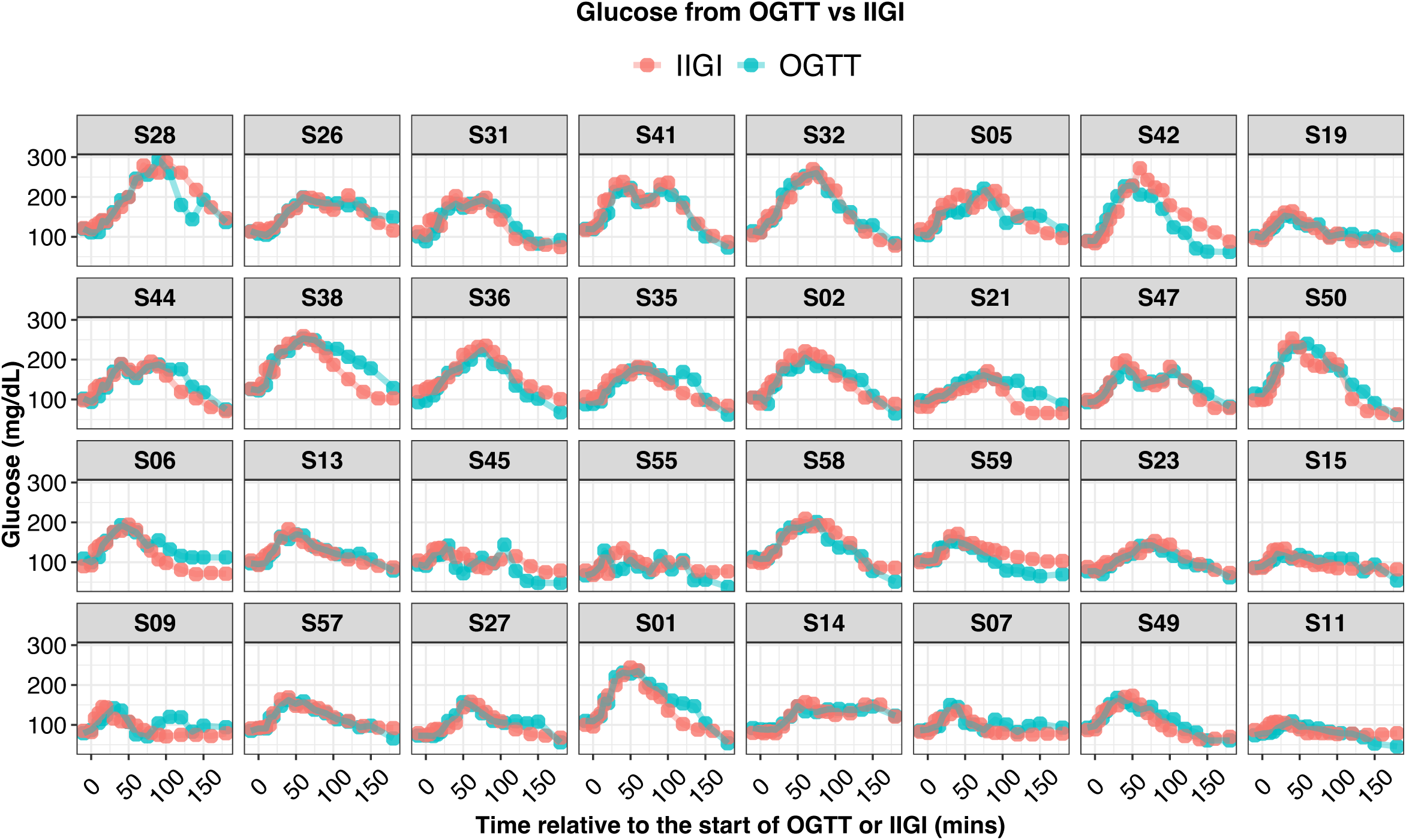
Concordance analysis of plasma glucose concentration during OGTT and IIGI. Glucose time-series from OGTT and IIGI are overlaid on each other, showing their high similarity. Each box represents a participant, the x-axis represents time in minutes relative to the start of OGTT or IIGI, and the y-axis represents glucose concentration in mg/dL. Average Pearson correlations between each individual’s glucose time-series from OGTT and IIGI equals 0.82, with a standard deviation equal 0.16.

**Figure S4:**
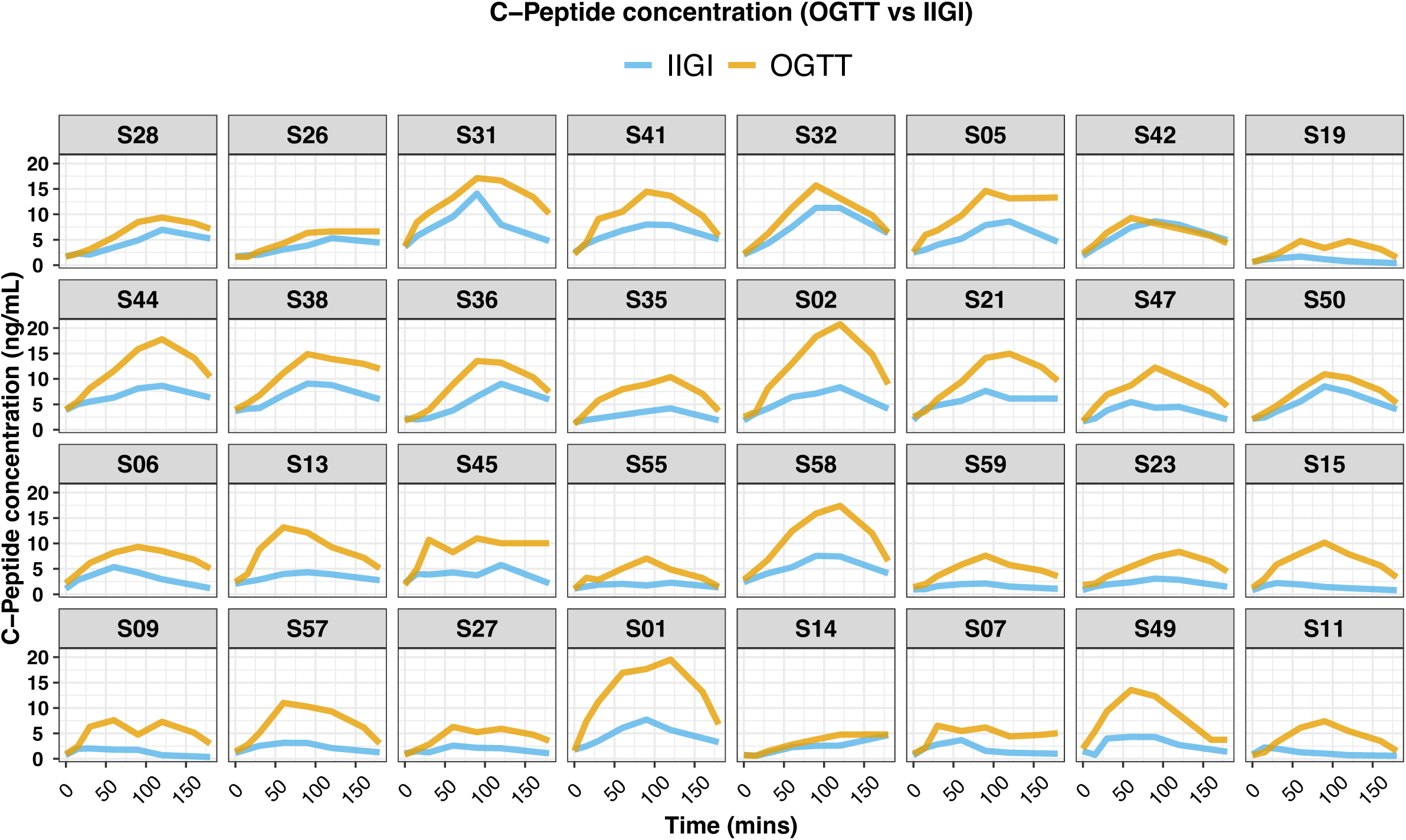
Plasma C-peptide concentration during OGTT and IIGI. C-peptide concentrations were obtained at seven timepoints (0, 15, 30, 60, 90, 120, and 180 min). Each box represents a participant, the x-axis represents time in minutes relative to the start of OGTT or IIGI, and the y-axis represents C-peptide concentration in ng/mL. The area between the two curves is calculated to infer the incretin effect.

**Figure S5:**
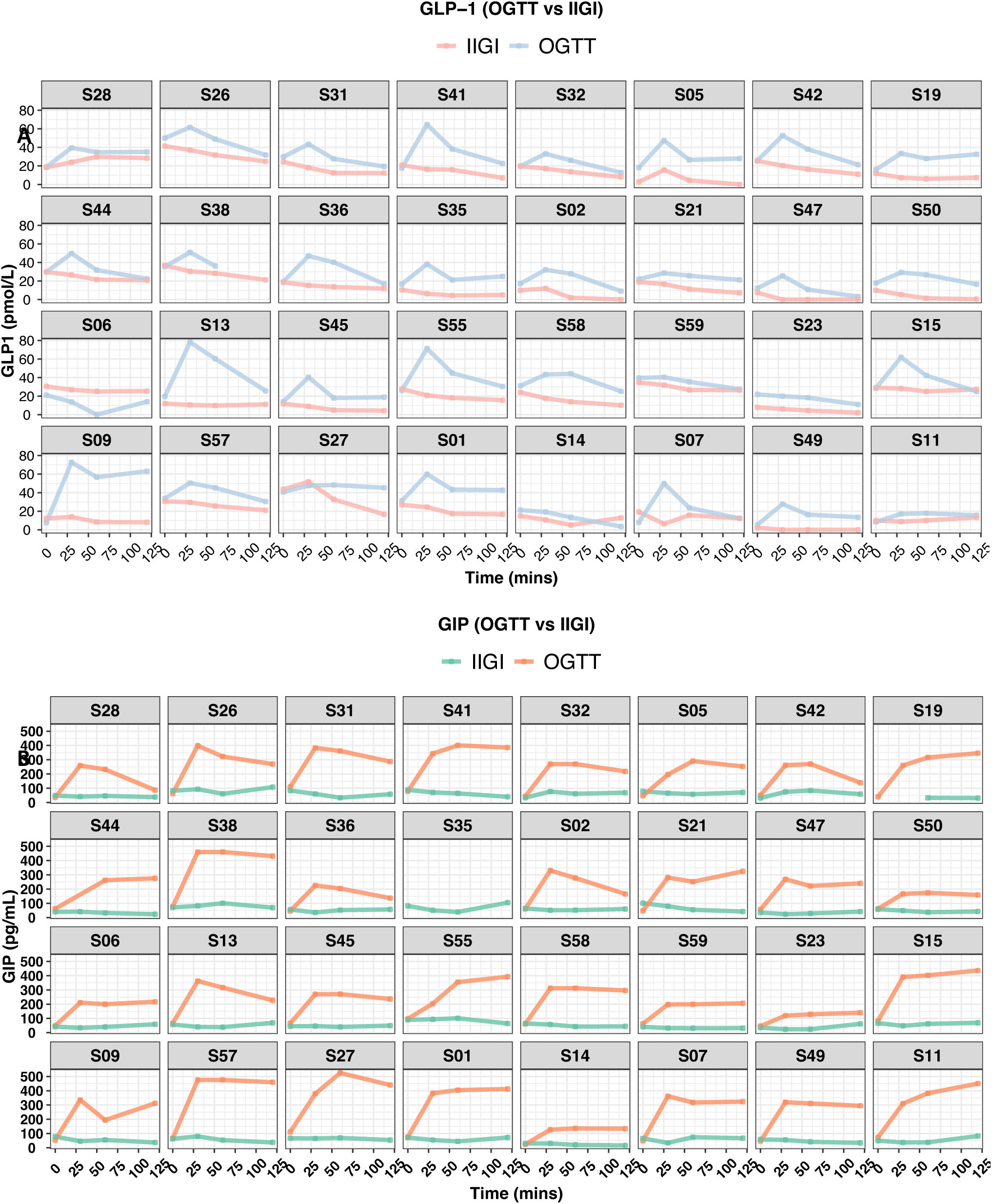
Total plasma Incretin hormones concentration during OGTT and IIGI. **(A)** GLP-1 concentration during OGTT and IIGI. **(B)** GIP concentration during OGTT and IIGI. The measurements were obtained at four timepoints (0, 30, 60, and 120 min). Subject S38 is missing the GLP-1 measurement at 120 min from OGTT. Subject S19 is missing GIP measurements at 0 and 30 min during IIGI. Subject S35 is missing GIP all measurements during IIGI. Each box represents a participant, the x-axis represents time in minutes relative to the start of OGTT or IIGI, and the y-axis represents GLP-1 or GIP concentration in pmol/L and pg/mL, respectively.

**Figure S6:**
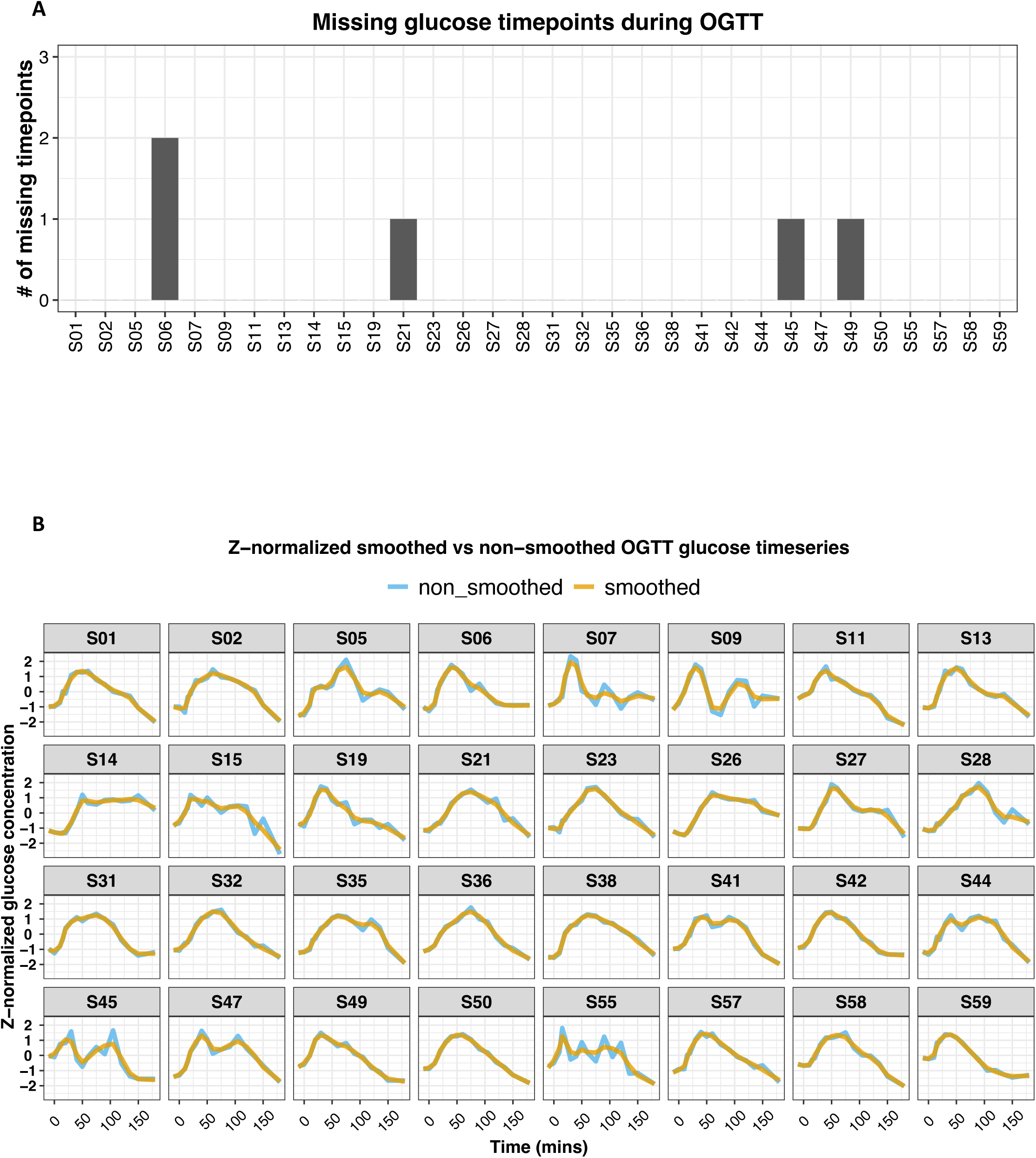
The effect of preprocessing steps employed on glucose time-series. **(A)** Shows the number of missing glucose timepoints for each participant. These timepoints were imputed using linear interpolation. As illustrated, 87.5% (28 out of 32 participants) of participants had complete time-series, 9.4% (3 out of 32 participants) were missing only one timepoint, and 3.1% (1 out of 32 participants) were missing 2 timepoints. **(B)** All smoothed glucose curves overlayed on top of the non-smoothed curve. As illustrated in the figure, smoothing has a minor effect on the curve in most cases. However, in cases where there are strong fluctuations (e.g., S55), smoothing helps capture the general pattern rather than those immediate spikes and valleys.

**Figure S7:**
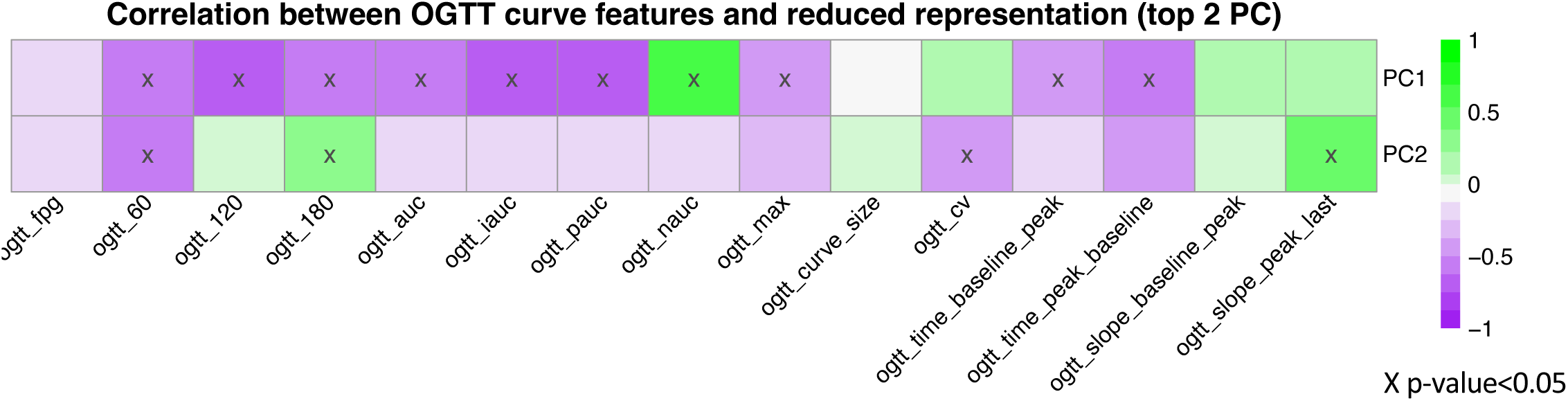
Relationship between the reduced representation of the glucose time-series and the extracted curve features. We calculated the correlations between the top 2 PCs (OGTT_G_ReducedRep) and the extracted features (OGTT_G_Features). Variances explained by PC1 and PC2 are 43.8% and 18%, respectively. As illustrated in the Figure, PC1 strongly negatively correlated with the incremental area under the curve (iAUC), and positively correlated with the negative area under the glucose curve (nAUC). This means, for example, that higher positive values on the PC1 are associated with lower iAUC.

**Figure S8:**
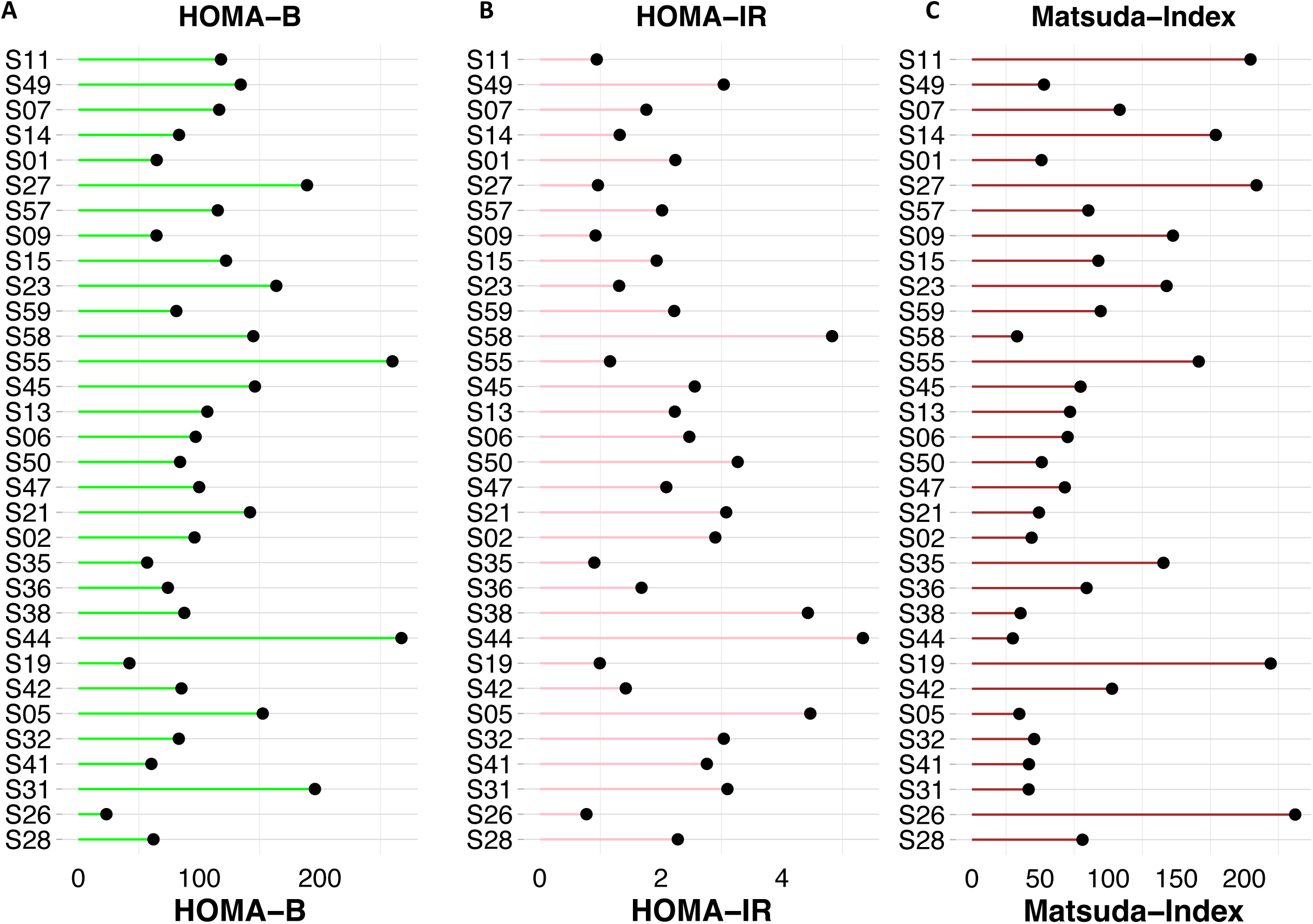
Common surrogate markers of insulin resistance and sensitivity measured on the initial cohort participants. **(A)** HOMA-B, **(B)** HOMA-IR, and **(C)** Matsuda Index.

**Figure S9:**
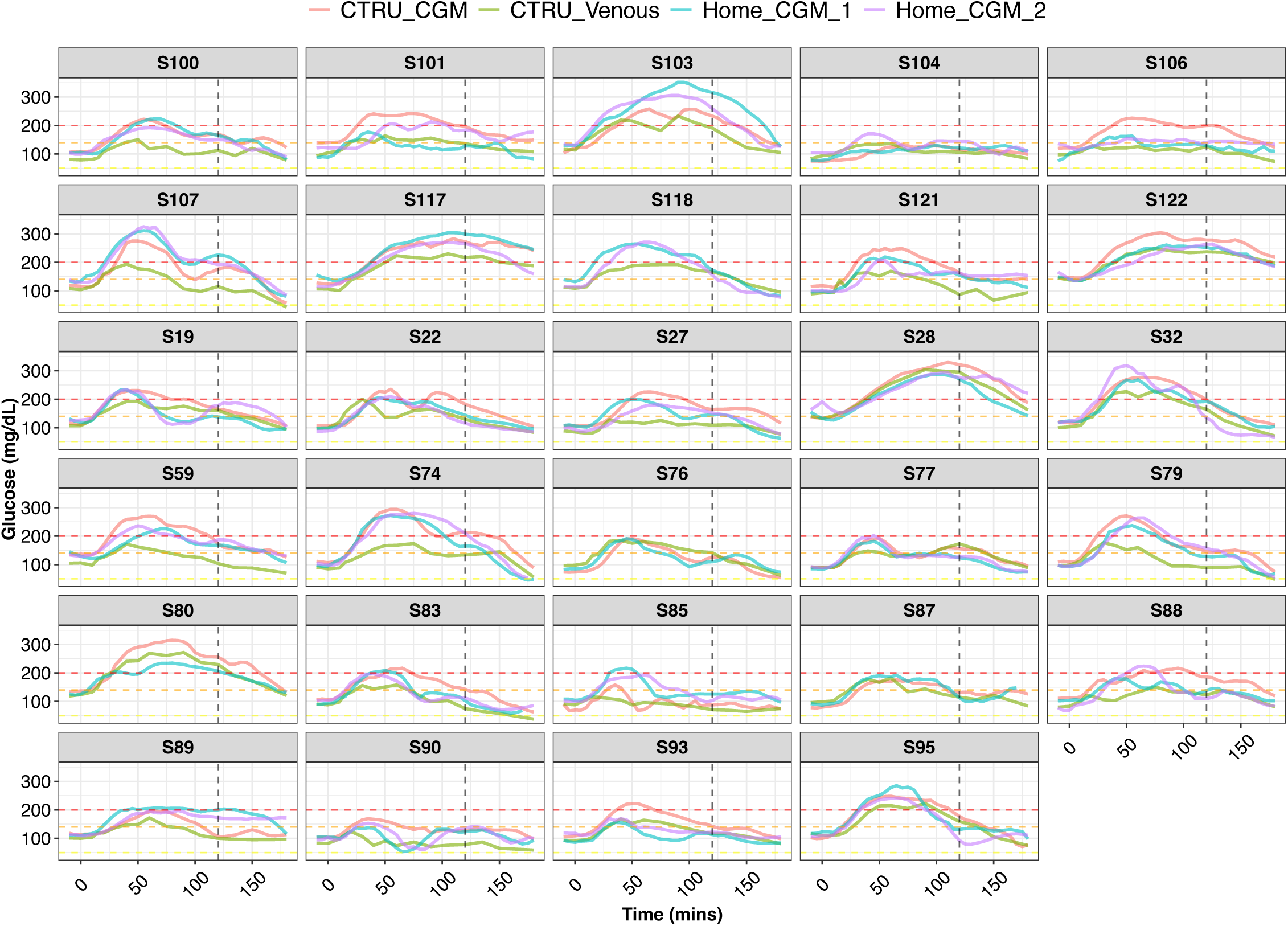
Glucose time-series from the validation and at-home CGM cohort. Each box represents one participant. Each participant has four glucose time-series; glucose time-series measured via venous blood sample during clinical OGTT (green), glucose time-series measured via CGM during the clinical OGTT (red), glucose time-series measured via CGM during 1st at-home testing trial (cyan), and glucose time-series measured via CGM during 2nd at-home testing trial (purple). Gold standard muscle insulin resistance is measured via SSPG, written on the header of each box.Time=0 on the x-axis represents the time when participants start drinking the glucose drink. The vertical black dashed line represents the time at 120 minutes when the glucose level is measured for clinical diagnosis of diabetes. A glucose value above 200 mg/dL (red dashed line) represents a diabetes status, a glucose value below 140 mg/dL (orange dashed line) represents a normoglycemic status, glucose value between 140-200 mg/dL represents a prediabetes status, and glucose value below 70 mg/dL (yellow dashed line) represents a hypoglycemia status.

## Summary Figure

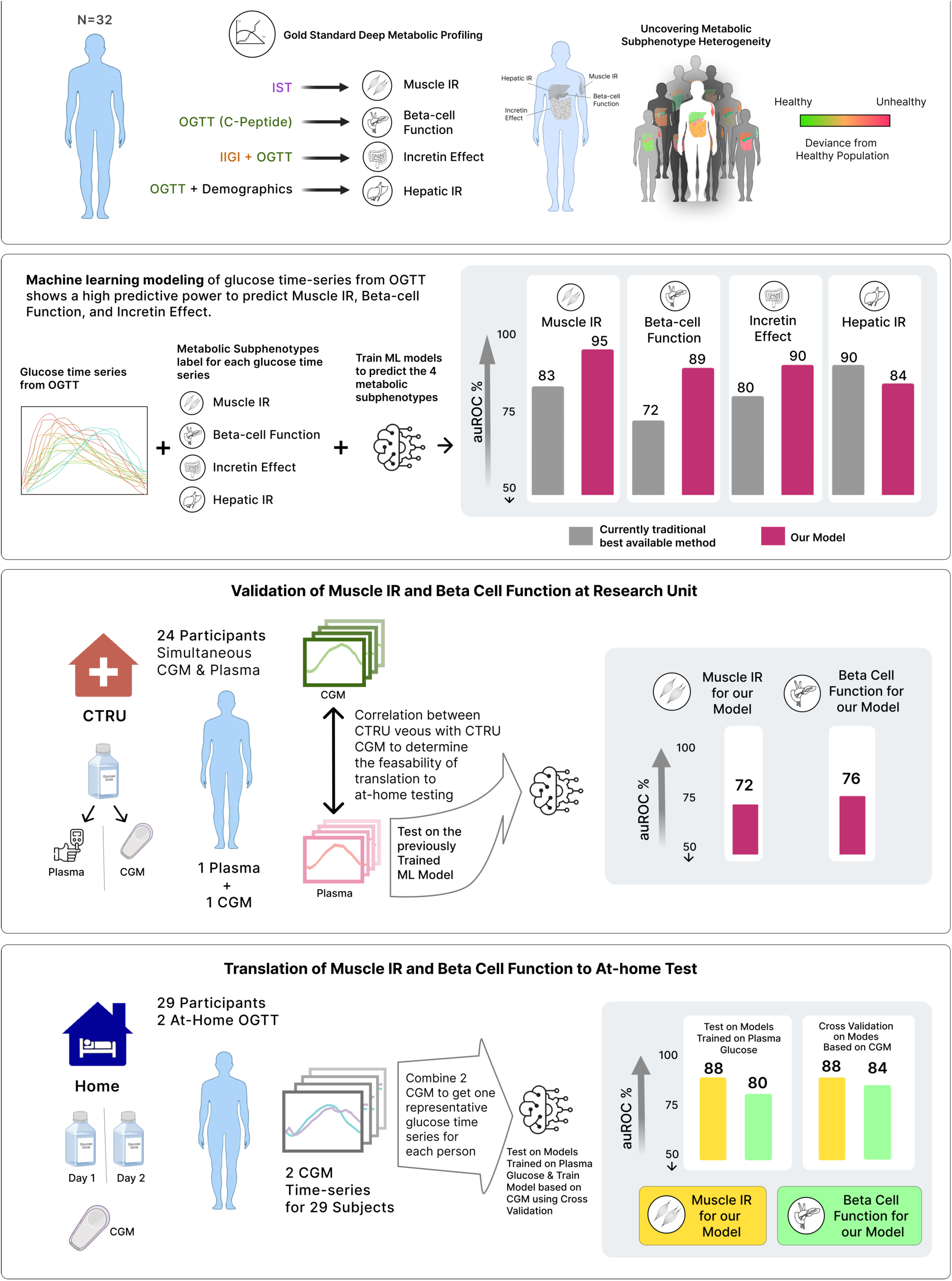

